# Genetically regulated molecular mediators of phenotypic heterogeneity in autism spectrum disorder

**DOI:** 10.64898/2026.10.18.26365834

**Authors:** Camilla Marinelli, Alessandro Amaolo, Davide Marnetto, Paolo Provero

## Abstract

**Background:** Autism spectrum disorder (ASD) shows wide phenotypic variation, from adults without intellectual disability and with low support needs to minimally verbal children with intellectual disability and severe behavioral symptoms. Genetic studies of this heterogeneity have focused mainly on high-impact de novo variants and polygenic scores; here we investigate, at the molecular level, the role of common variants.

**Materials and methods:** We analyzed genotypic and phenotypic data from 9,332 individuals with ASD in the SPARK cohort. We considered 31 traits: six latent factors derived from the Social Communication Questionnaire and the Repetitive Behavior Scale-Revised, 22 co-occurring diagnoses, parent-reported IQ range, age at diagnosis, and age at first walking. We performed within-case genome-wide association studies (GWAS) and, for the 10 traits with evidence of SNP heritability, association analyses with genetically predicted gene and protein expression, splicing, and enhancer activity in brain and blood (xWAS). Nominated genes were tested for enrichment in biological processes, brain cell types, and developmental stages. xWAS-derived transcriptional signatures were compared with chemical perturbation signatures from CMap/LINCS L1000 to prioritize candidate drugs.

**Results:** GWAS identified seven genome-wide significant loci for five traits, including loci near *GABRG3* and *TENM2* associated with co-occurring sleep, feeding or toileting problems. xWAS identified seven gene–trait associations involving three phenotypes (FDR < 0.1), including *CACNA1G* with communication skills and *NEGR1* with learning disability. GWAS and xWAS nominated different genes which however converged on synaptic transmission and gated channel activity, with preferential expression in neurons during later stages of brain development. PARP and p38 MAPK inhibitors were predicted to reverse the signatures of both social-communication and repetitive-behavior phenotypes, whereas several compound classes, including histone deacetylase inhibitors and mTOR inhibitors, were predicted to reverse the signatures of some phenotypes while mimicking those of others.

**Conclusion:** Combining within-case GWAS with xWAS nominates candidate genes mediating common-variant effects on ASD heterogeneity, provides biological insight, and suggests candidates for phenotypically stratified drug repositioning.

## Introduction

Autism spectrum disorder (ASD) is characterized by strong heritability, due to both rare and common variants, and wide phenotypic heterogeneity [1,2]. Recent studies indicate that such heterogeneity is itself partly heritable, with an important role played by common variants, as shown in particular by the patterns of correlation of polygenic scores with core and associated autism features [3] and with classes of patients grouped by phenotypic profiles [4]. However, the specific molecular features mediating the effects of common variants on such heterogeneity have not been systematically mapped.

Mapping of these molecular features is a necessary step towards understanding the biological underpinnings of heterogeneity and progressing towards stratified therapeutic approaches. Here, we applied statistical genetics methods to 9,332 individuals with ASD with available genotype and rich phenotypic data from the SPARK cohort [5], aiming to identify genetic variants and molecular features associated with phenotypic traits derived from behavioral questionnaires, co-occurring diagnoses, and other quantitative assessments.

We combined within-case genome-wide association studies (GWAS) with approaches based on genetically regulated molecular features, including gene and protein expression, splicing, and enhancer activity inferred from eRNA expression (collectively referred to as xWAS in the following). Compared with GWAS, xWAS methods aggregate weak variant-level signals and produce signed estimates of effect, thus potentially increasing power and allowing a more direct route to downstream functional analyses. While a GWAS of phenotypic heterogeneity in ASD has been published [6], to the best of our knowledge, all published xWAS of ASD have targeted diagnostic status, using case-control or family-based designs. [7–10].

Besides nominating individual genes whose dysregulation is associated with specific phenotypes within individuals with ASD, we performed several enrichment analyses to identify biological processes, cell types, and brain developmental stages associated with these genes. In particular, xWAS yields phenotype-associated transcriptional signatures that can be compared with chemical perturbation data to prioritize compounds predicted to reverse them, providing candidates for stratified drug repositioning in ASD. The overall analytical strategy of the work is shown schematically in Fig. 1.

**Fig. 1.**
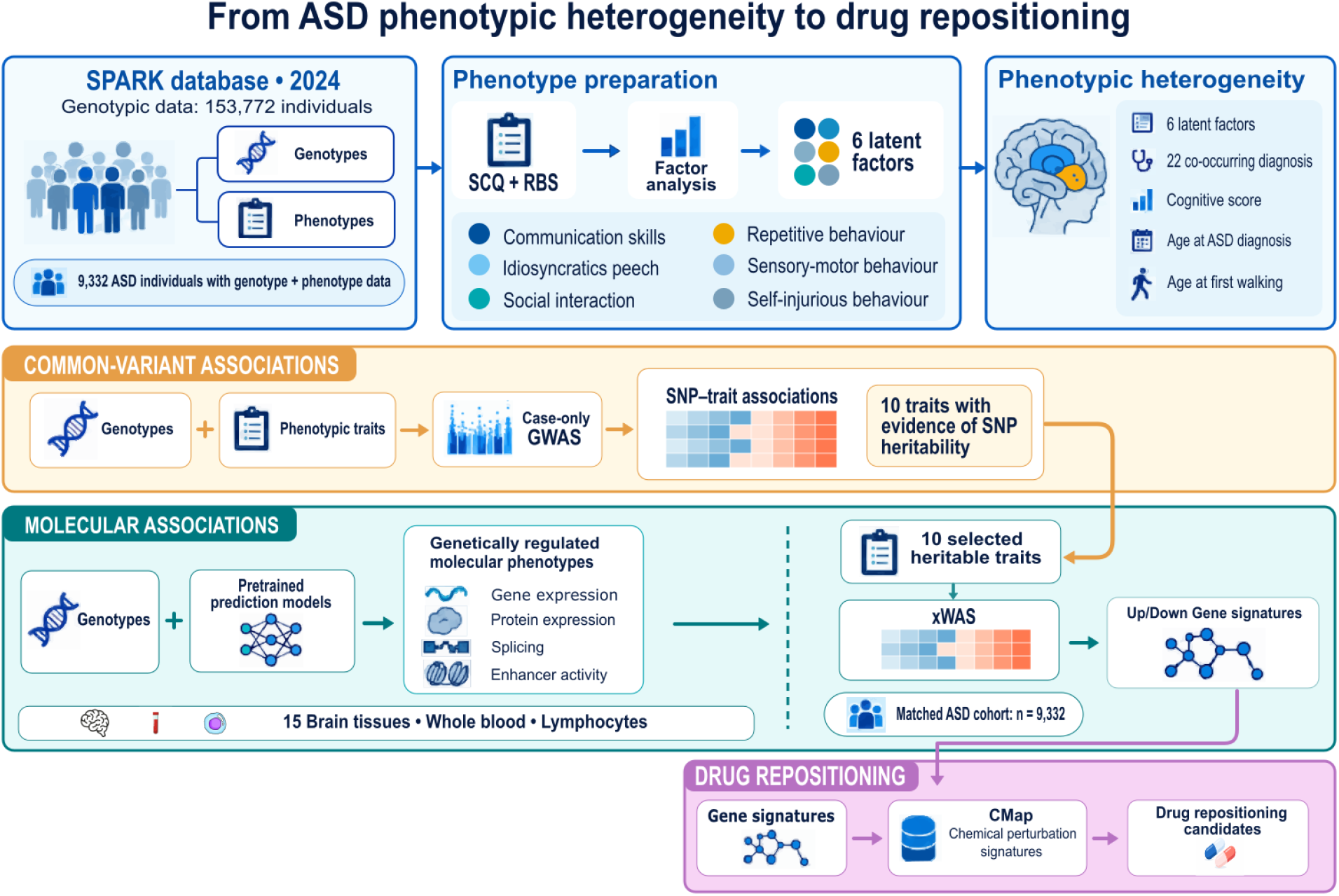
Graphical representation of the analytical workflow

## Results and Discussion

### Factor analysis reveals six latent phenotypic traits in ASD

In SPARK, phenotypic manifestations of ASD are mostly measured through questionnaires, including the Repetitive Behavior Scale - Revised (RBS-R) [11] and the Social Communication Questionnaire (SCQ) - Lifetime version [12]. We performed factor analysis (see Methods) on these questionnaires, using 23,939 individuals with ASD (all individuals with ASD and phenotypic data were included in the factor analysis irrespective of the availability of genetic data). The best model included 6 factors, as in [3], and the item loadings supported a similar interpretation: Four non-social factors (idiosyncratic speech, repetitive behavior, self-injurious behavior, and sensory-motor behavior) and two social ones (communication skills and social interaction). We chose the sign of the factors so that all of them had negative correlation with the parent-reported IQ range, so that positive scores always correlate with greater difficulty. This IQ measure (henceforth “cognitive test score”) was chosen because it is available for a larger subset of the cohort than the full-scale IQ from clinical records, and has been shown to agree well with standardized measures of IQ range [13]. The four non-social factors were positively correlated with each other (Pearson’s *r* 0.32 – 0.61, Fig. 2) while the social ones showed weak correlation with each other (r = 0.11), and with non-social factors (r -0.14 to 0.25).

**Fig. 2:**
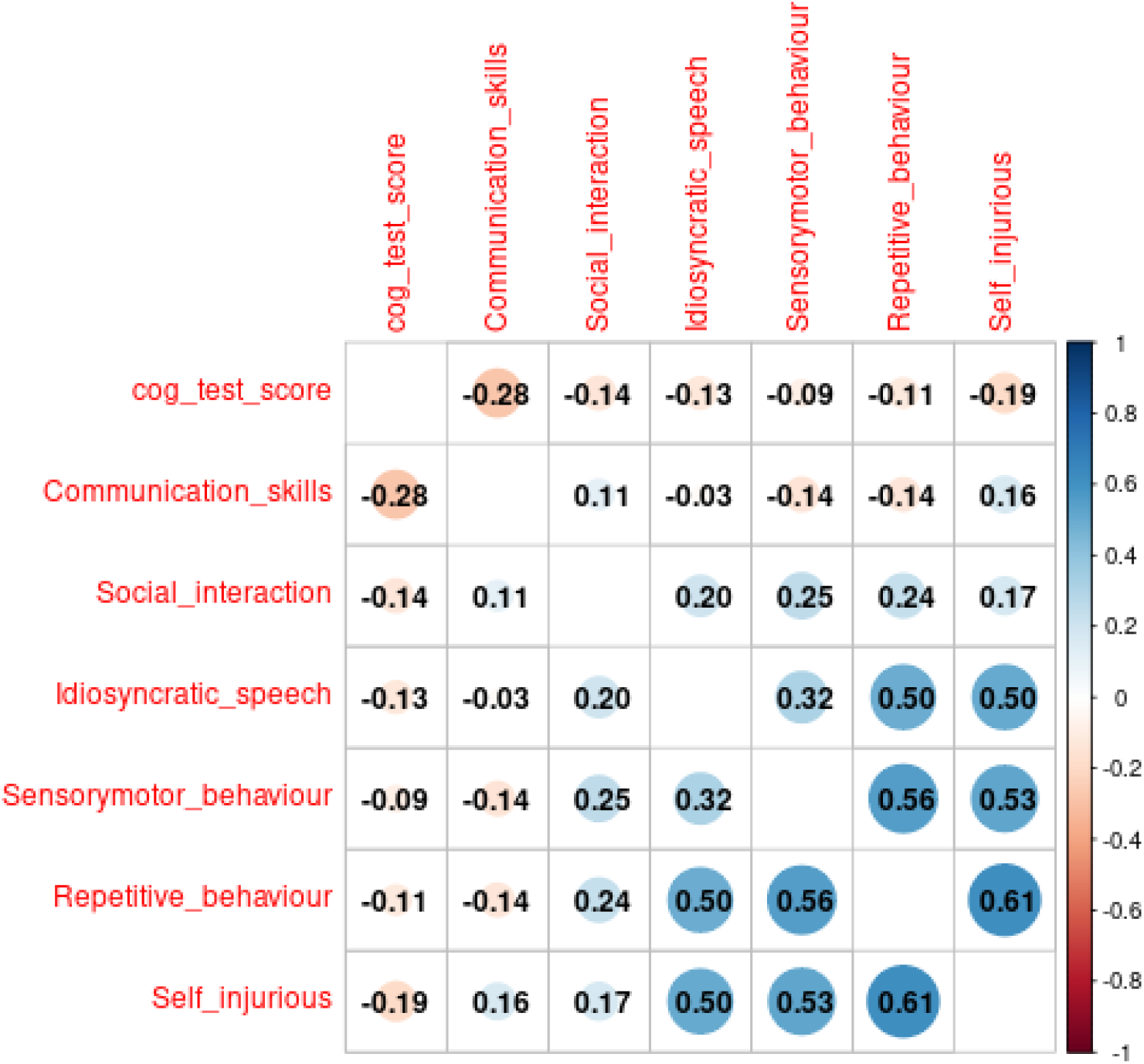
Correlation of the six identified factors with each other and with the cognitive test score.

### A within-case genome-wide association study identifies candidate loci associated with phenotypic heterogeneity

We set out to investigate the genetic basis of phenotypic heterogeneity at the molecular level, considering the six factors described above and 25 additional phenotypes, including cognitive test score, age at diagnosis, age at first walking, and 22 co-occurring diagnoses (see Methods). We first conducted within-case genome-wide association studies of these phenotypes in the 9,332 individuals with ASD for whom genotyping data were available, after excluding close relatives. We found a total of 7 genome-wide significant loci (P < 5e-8) for 5 phenotypes, shown in Tab. 1 and Fig. 3. FUMA [14] was used to assign candidate effector genes to the loci. Note that these associations pass the conventional per-trait genome-wide threshold but not a study-wide Bonferroni correction, and therefore should be considered suggestive results in need of replication in an independent cohort.

**Fig. 3:**
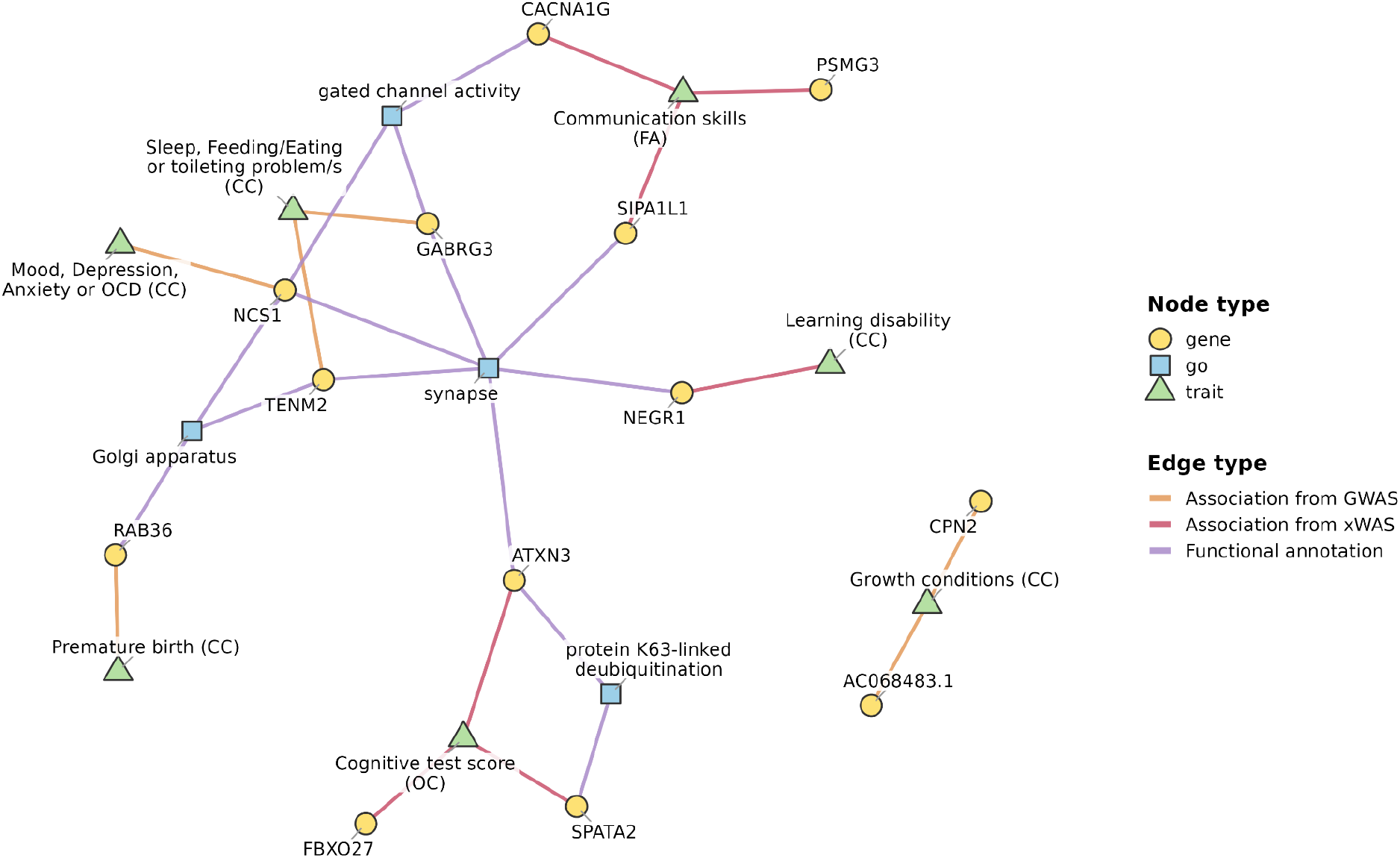
Gene-phenotype associations found by GWAS or xWAS, and Gene Ontology enrichment. GO terms enriched (FDR < 0.05) among the GWAS genes, the xWAS genes, or their union are shown. CC: co-occurring condition; FA: factor; OC: other continuous phenotype.

**Tab. 1.** Genome-wide significant loci. CC: co-occurring condition. Coordinates are GRCh37.

| phenotype | lead SNP | chr | pos | MAF | P | nearest gene | FUMA genes |
| --- | --- | --- | --- | --- | --- | --- | --- |
| Growth conditions (CC) | rs114844402 | 2 | 129811704 | 0.02821 | 4.765e-08 | AC068483.1 |  |
| Growth conditions (CC) | rs6783012 | 3 | 194060294 | 0.198 | 1.61e-08 | CPN2 | CPN2; GP5 |
| Mood, Depression, Anxiety or OCD (CC) | rs10819610 | 9 | 132946615 | 0.3054 | 4.967e-08 | NCS1 | EXOSC2; NCS1 |
| Premature birth (CC) | rs16998069 | 22 | 23494926 | 0.01782 | 3.078e-08 | RAB36 | GNAZ;<br>LL22NC03-63E9.3<br>; PPM1F; PRAME;<br>RAB36; RTDR1;<br>YPEL1; ZNF280A;<br>ZNF280B |
| Sleep, Feeding/Eating or toileting problem/s (CC) | rs66830482 | 5 | 167589028 | 0.3063 | 4.304e-08 | TENM2:CTB-178<br>M22.1 | TENM2 |
| Sleep, Feeding/Eating or toileting problem/s (CC) | rs9972311 | 15 | 27459234 | 0.3399 | 2.281e-08 | GABRG3 | GABRG3 |
| Speech and language, intellectual disability (CC) | rs72625659 | 14 | 65491312 | 0.07241 | 3.571e-08 | CHURC1-FNTB:<br>FNTB:MAX | CHURC1-FNTB;<br>CTD-2014B16.3;<br>FNTB; GPX2;<br>MAX; RAB15;<br>RDH11; RDH12 |

A locus on chromosome 22 was found associated with premature birth: As the phenotypic data do not distinguish between spontaneous and medically induced pre-term birth, the interpretation of this finding is difficult. Moreover, the association might reflect maternal rather than fetal effects. The locus on chromosome 14 associated with speech and language, intellectual disability/cognitive impairment coincides with a de novo microdeletion found in a patient with autism and spherocytosis [15], although the genes associated with it by FUMA do not suggest an immediate mechanism.

One of the loci associated with growth conditions is located in the 3q29 region, ∼1.7 Mb from a recurrent deletion interval known to confer a large increase in autism risk [16]. A variant located ∼ 1500 bp from our lead variant was found associated with phenotypic heterogeneity in ASD in [6], although the associated phenotype was quite different and related to social interaction. Another associated locus lies in a gene desert in chromosome 2 and no relevant genes are identified by FUMA.

The locus on chromosome 9 associated with mood, depression, anxiety or OCD contains the *NCS1* gene as a plausible mechanistic candidate. *NCS1* is a neuronal calcium sensor involved in synaptic transmission and plasticity [17]. A rare missense variant in NCS1 was reported in an individual with ASD [18], while *NCS1* deficiency was reported to cause anxiety and depressive-like behavior in mice [19], although this finding was not replicated in a later study [20].

Two loci were associated with the phenotype “Sleep, Feeding/Eating or toileting problem/s”. For the first one, in chromosome 5, FUMA highlighted a single gene, *TENM2*, involved in synaptic transmission and specifically in the balance of excitatory and inhibitory synapse formation [21]. The second locus, in chromosome 15, points to the *GABRG3* gene, encoding a GABA receptor, thus again suggesting a possible effect on the excitatory/inhibitory balance. Notably, this locus is within the region involved in the 15q11.2-13.1 duplication (Dup15q) syndrome, one of the most common CNVs associated with neurodevelopmental disorders, with high penetrance for ASD, and associated with abnormal sleep physiology [22].

Taken together, our GWAS identified seven loci associated with specific phenotypic dimensions within ASD at the conventional genome-wide threshold. Although these associations require replication, several point to biologically plausible candidate genes. In particular, the candidate genes at the mood/anxiety and sleep/feeding/toileting loci (*NCS1*, *TENM2*, *GABRG3*) are all involved in synaptic function, including the balance of excitatory and inhibitory transmission.

### Genetically regulated molecular features are associated with phenotypic traits

While genome-wide significant hits were found for only 5 of the 31 phenotypes, none of which were questionnaire-derived factors, LDSC analysis [23] of the GWAS results suggested that 10 phenotypes (Tab. 2), including the repetitive behavior, self-injurious behavior, and communication skills factors, showed at least some evidence of SNP heritability (Z(h²) > 1.5; henceforth “heritable” for simplicity). Notably, only one phenotype (premature birth) among those with genome-wide significant hits was heritable. The lack of detectable heritability for the other phenotypes with GWAS hits does not necessarily argue against the GWAS findings: All four are binary co-occurring conditions, for which LDSC has limited power given the number of cases, and a single locus of moderate effect may contribute little to a genome-wide h² estimate. Conversely, the pattern observed for the remaining heritable phenotypes is consistent with a polygenic architecture of many weak effects, which GWAS is underpowered to detect in a cohort of this size. We thus reasoned that an approach based on intermediate molecular phenotypes could usefully complement the GWAS in identifying the molecular mediators of this heritability.

**Tab. 2:** Heritable phenotypes. CC: co-occurring condition; FA: factor; OC: other continuous phenotype.

| Phenotype | Category | Abbreviation | $h^2$ | SE | z |
| --- | --- | --- | --- | --- | --- |
| Age at diagnosis | OC | diagnosis_age | 0.17 | 0.053 | 3.23 |
| Cognitive test score | OC | cog_test_score | 0.28 | 0.11 | 2.81 |
| Age at first walking | OC | walked_age_months | 0.14 | 0.049 | 2.78 |
| Premature birth | CC | birth_prem | 0.095 | 0.04 | 2.25 |
| Repetitive behaviour | FA | repetitive_behaviour | 0.095 | 0.042 | 2.25 |
| Learning disability | CC | dev_ld | 0.086 | 0.043 | 1.99 |
| Visual or auditory impairments | CC | med_cond_visaud | 0.080 | 0.045 | 1.76 |
| Self-injurious behaviour | FA | self_injurious | 0.079 | 0.045 | 1.74 |
| ADHD or ADD | CC | behav_adhd | 0.074 | 0.046 | 1.59 |
| Communication skills | FA | communication_skills | 0.075 | 0.048 | 1.57 |

We computed the genetic component of several molecular features in the same individuals used in the GWAS analysis. The molecular features included gene expression and splicing in thirteen GTEx [24] brain tissues plus whole blood and EBV-transformed lymphocytes; gene expression in the prefrontal cortex from the PsychENCODE project [25]; enhancer activity, as measured by enhancer RNA (eRNA) expression in the same GTEx tissues [26]; protein expression in the dorsolateral prefrontal cortex [27] and in the plasma [28,29]. In the following we refer to these analyses together as xWAS analyses.

We define a genetically regulated feature (GRF) as a molecular feature/tissue pair that could be predicted in our subjects using one of several published predictive models based on reference data (JTI for gene expression [30] and enhancer activity [31]; PrediXcan [32–34] for splicing; FUSION [27,35] for brain proteomics; and polygenic scores [36,37] for plasma proteomics). A total of ∼270,000 GRFs could be predicted. We then looked for significant associations between each GRF and phenotypic measures, using linear or logistic regression depending on the nature of the phenotype, and including age at registration, sex, and 10 genomic PCs as covariates. This analysis was limited to the 10 phenotypes showing evidence of SNP-heritability. Given the large heterogeneity of the number of features predicted for each feature type (from ∼ 3,500 plasma proteins to ∼ 125,000 expression GRFs), we applied the Benjamini-Hochberg correction separately for each phenotype and feature type, but across all tissues.

At 10% FDR we found 22 significant associations involving 3 phenotypes (Suppl. Tab. 1). As several GRFs are highly correlated with each other (for example, different introns of the same gene, or the expression of the same gene in different tissues, or different genes sharing eQTLs), we organized them into approximately independent “loci”, each containing a set of highly correlated GRFs (see Methods). The significant associations involved 7 such loci, and are shown in Tab. 3.

**Tab. 3:**
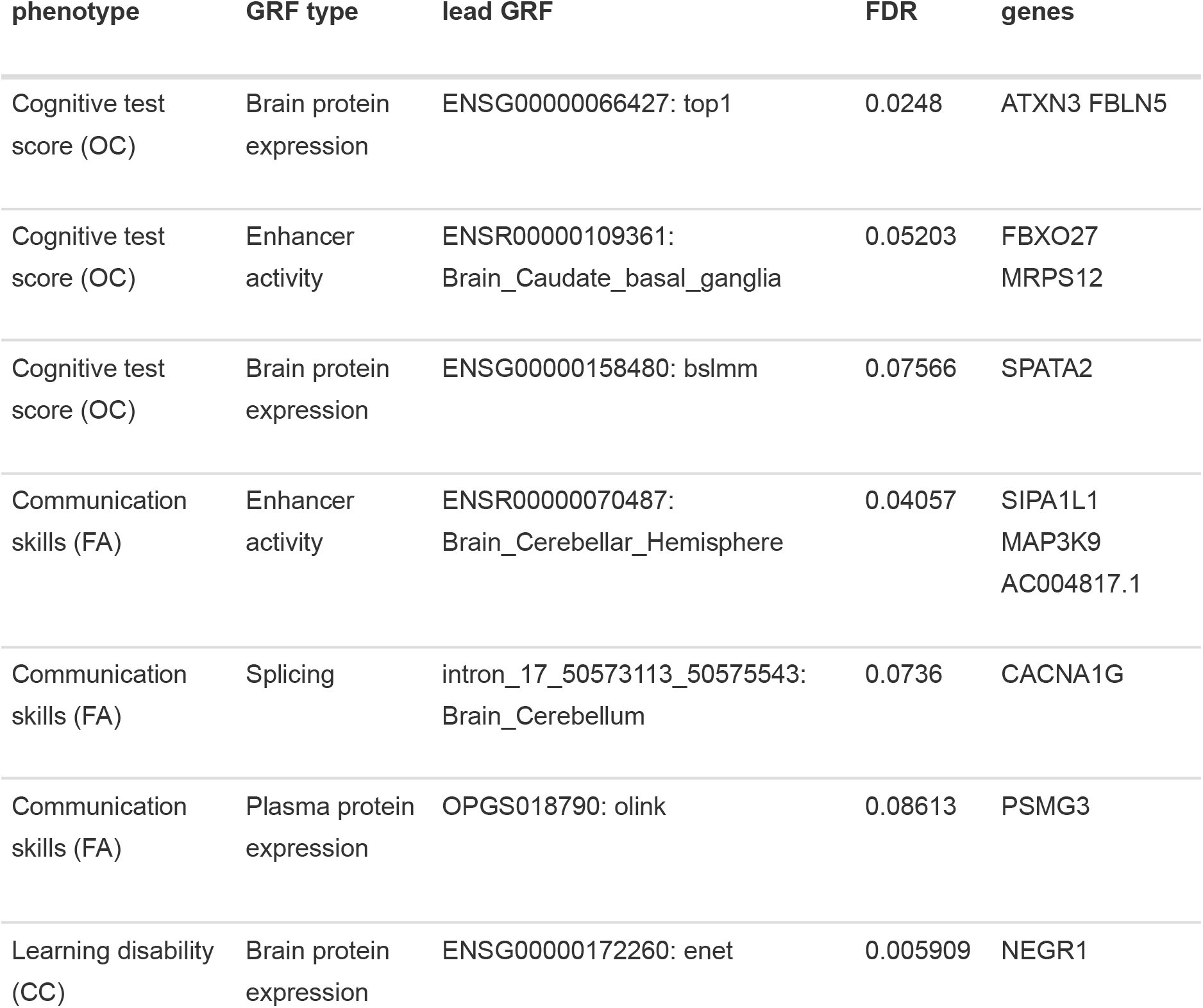
Associations between molecular features and phenotypes, at FDR < 0.1. For each association, the lead GRF is the GRF in the locus with the strongest association P-value. A GRF is defined by a molecular feature (gene, protein, intron, or enhancer) and a tissue (for gene expression enhancer activity, and splicing); a FUSION predictive model (for brain proteomics); or a proteomics platform (for plasma proteomics). To each enhancer activity GRF we associate the gene whose expression has the highest correlation with the activity of the enhancer in the relevant tissue. In the “genes” column we report the genes associated with all the significant GRFs within the locus. CC: co-occurring condition; FA: factor; OC: other continuous phenotype.

| phenotype | GRF type | lead GRF | FDR | genes |
| --- | --- | --- | --- | --- |
| Cognitive test score (OC) | Brain protein expression | ENSG00000066427: top1 | 0.0248 | ATXN3 FBLN5 |
| Cognitive test score (OC) | Enhancer activity | ENSR00000109361: Brain_Caudate_basal_ganglia | 0.05203 | FBXO27 MRPS12 |
| Cognitive test score (OC) | Brain protein expression | ENSG00000158480: bslmm | 0.07566 | SPATA2 |
| Communication skills (FA) | Enhancer activity | ENSR00000070487: Brain_Cerebellar_Hemisphere | 0.04057 | SIPA1L1 MAP3K9 AC004817.1 |
| Communication skills (FA) | Splicing | intron_17_50573113_50575543: Brain_Cerebellum | 0.0736 | CACNA1G |
| Communication skills (FA) | Plasma protein expression | OPGS018790: olink | 0.08613 | PSMG3 |
| Learning disability (CC) | Brain protein expression | ENSG00000172260: enet | 0.005909 | NEGR1 |

#### Cognitive test score

Three loci were associated with the cognitive test score. In the first one the leading GRF is the predicted brain protein level of ATXN3, although the predicted level of FBLN5 is also significantly associated. The two genes are located on chromosome 14 at a distance of ∼100 Kb, and their genetically regulated protein expressions are highly correlated (Pearson correlation between 0.54 and 0.85 depending on the predictive model). *ATXN3* is a plausible mechanistic candidate, given its roles in autism-related pathways such as mTORC1 signaling [38,39] and histone deacetylation [40,41].

The second locus involves the activity of an enhancer on chromosome 19 and the brain protein levels of MRPS12. From the enhancer-gene associations provided by [26], the activity of the enhancer is strongly correlated with the expression of many neighboring genes, including *FBXO27* and *MRPS12. FBXO27* is involved in the ubiquitin-proteasome system (UPS), while *MRPS12* is a mitochondrial ribosomal protein. Disruption of both the UPS and mitochondria have been associated with ASD [42,43], thus both genes represent plausible mechanistic candidates.

The third locus involves the brain protein level of SPATA2. This protein is involved in immune response by regulating TNF-induced NF-κB signalling, and common variants in the same locus were found associated with autistic traits in the general population [44].

#### Communication skills

Three loci were found associated with the communication skills factor. The first one involves the activity of three enhancers whose most strongly associated gene is *SIPA1L1*, a postsynaptic Rap GTPase-activating protein that regulates dendritic spine morphology [45]. S*IPA1L1* is also known to be involved in GPCR signaling, with strong behavioral effects in mice [46]. GPCR dysregulation has been recently suggested to be a convergent pathological mechanism in ASD, with 15% of SFARI genes being involved in GPCR activity and signalling processes [47].

The second locus involves the splicing of *CACNA1G*, which encodes Cav3.1, a T-type calcium channel expressed in cerebellar, thalamic, and cortical neurons, thus adding to the evidence for synaptic and neuronal-excitability mechanisms suggested by the GWAS results. The fact that this association comes from splicing data is particularly intriguing, since *CACNA1G* splicing has been shown to be strongly regulated during brain development [48]. The cerebellar origin of the signal is also notable, given that *CACNA1G* variants have been associated with childhood-onset cerebellar atrophy [49].

The third locus involves plasma levels of PSMG3, a proteasome assembly chaperone, and is more difficult to interpret mechanistically. *PSMG3* lies in the 7p22.3 region, whose microdeletion causes a complex developmental phenotype [50], and the signal may reflect other genes in the region.

#### Learning disability

One locus was associated with co-occurring learning disability, the associated GRFs being the brain protein levels of NEGR1, whose deletions have been associated, in particular, with intellectual disability [51,52]. The gene is involved in GABAergic signaling [53] and specifically in ASD-related behavioral patterns [54]. Notably, while these studies analyze the effects of *NEGR1* deletion or downregulation, in our analysis the association coefficient is positive, suggesting that, in individuals with ASD, higher genetically predicted NEGR1 protein levels are associated with a higher probability of co-occurring learning disability. Recent experimental work in a mouse model [55] demonstrated that *NEGR1* overexpression in the hippocampus and prefrontal cortex leads to decreased working memory as well as depression and anxiety-like phenotypes through the impairment of synaptic function and myelination.

These data show that xWAS identifies associations for phenotypes not captured by GWAS, several of which again point to synaptic function. These qualitative considerations can be made quantitative through enrichment analyses, described below.

### Enrichment analyses

#### Gene Ontology

We performed Gene Ontology (GO) enrichment analysis on the lists of genes identified by GWAS, by xWAS, and their union, after choosing a single gene per locus with the data-driven criteria described in the Methods. These resulted in five protein-coding genes from GWAS (*CPN2*, *GABRG3*, *NCS1*, *RAB36*, and *TENM2*) and seven from xWAS (*ATXN3*, *CACNA1G*, *FBXO27*, *NEGR1*, *PSMG3*, *SIPA1L1*, and *SPATA2*). The GWAS loci on chromosomes 2 and 14 could not be associated with a protein-coding gene. The whole genome was used as background when analyzing GWAS genes, and the genes associated with at least one GRF when analyzing xWAS results.

The gene set obtained from the union of GWAS and xWAS genes showed significant enrichment (FDR < 0.05) for 24 GO terms. Here we used as background the whole genome, but the enriched terms are unchanged using the xWAS-appropriate background. Five of these terms reached nominal significance in both the GWAS and xWAS lists, including “synapse” and related terms. Selected non-redundant terms are shown in Fig. 3, while complete results are in Suppl. Table 2. These enrichments should be taken with some caution because they are based on a few genes per GO term, and the identification of a single gene per locus carries some uncertainty. Moreover, positional assignment of GWAS genes may favor long genes, which are overrepresented among synaptic genes. However, they suggest that GWAS and xWAS, while identifying different genes involved in different aspects of phenotypic heterogeneity, converge on synaptic genes.

To investigate the functional enrichment of the genes associated with individual phenotypes we considered, for each of them, all xWAS-associated genes with a suggestive P-value < 0.001, after collapsing the features into approximately independent loci and choosing within each locus the gene associated with the lead GRF. Again, the genes associated with at least one GRF were used as background. Although the permissive cutoff likely includes many false positive in these gene sets, the analysis showed significant functional enrichment (FDR < 0.05) for several phenotypes (Suppl. Tab. [suptab:GO]), pointing in particular to the role of microtubules and the cytoskeleton, with “spectrin-associated cytoskeleton” enriched in genes associated with co-occurring ADHD; “regulation of microtubule polymerization” and “regulation of cytoskeleton organization” associated with visual or auditory impairments; and “kinesin binding” associated with the communication skills factor. Importantly, the individual genes contributing to these enrichments for different phenotypes do not overlap, indicating that distinct genes affecting a shared cellular compartment contribute to different phenotypes. The role of the cytoskeleton in ASD has been recently reviewed in [56].

#### Brain cell types

A limit of the xWAS analyses we presented is that the GRF predictive models are based on bulk omics data, and thus do not clarify the cell types involved in the dysregulation associated with the phenotypes. To partly overcome this limitation, we investigated the enrichment of genes expressed in brain cell types among our associations using EWCE [57] and the single cell brain expression atlas of [58]. The tool allows enrichment analysis at two levels of granularity, to which we added a third, more coarse-grained one with eight major cell types (Suppl. Tab. 3). In this classification neurons were classified into excitatory, inhibitory, or other neurons based on the expressed neurotransmitters (see Methods), with the “other neurons” class containing neurons expressing a mix or neuromodulatory neurotransmitter profiles, including glycine, VGLUT3, serotonin, and acetylcholine.

When analyzing the gene lists produced by GWAS, xWAS, and their union, neurons were consistently enriched (Fig. 4A), as expected from the Gene Ontology results, reaching nominal significance in all three gene lists and corrected significance (FDR < 0.05) for interneurons in the combined list (in which “excitatory neurons” and “other neurons” are both suggestively enriched (FDR = 0.085)).

**Fig. 4:**
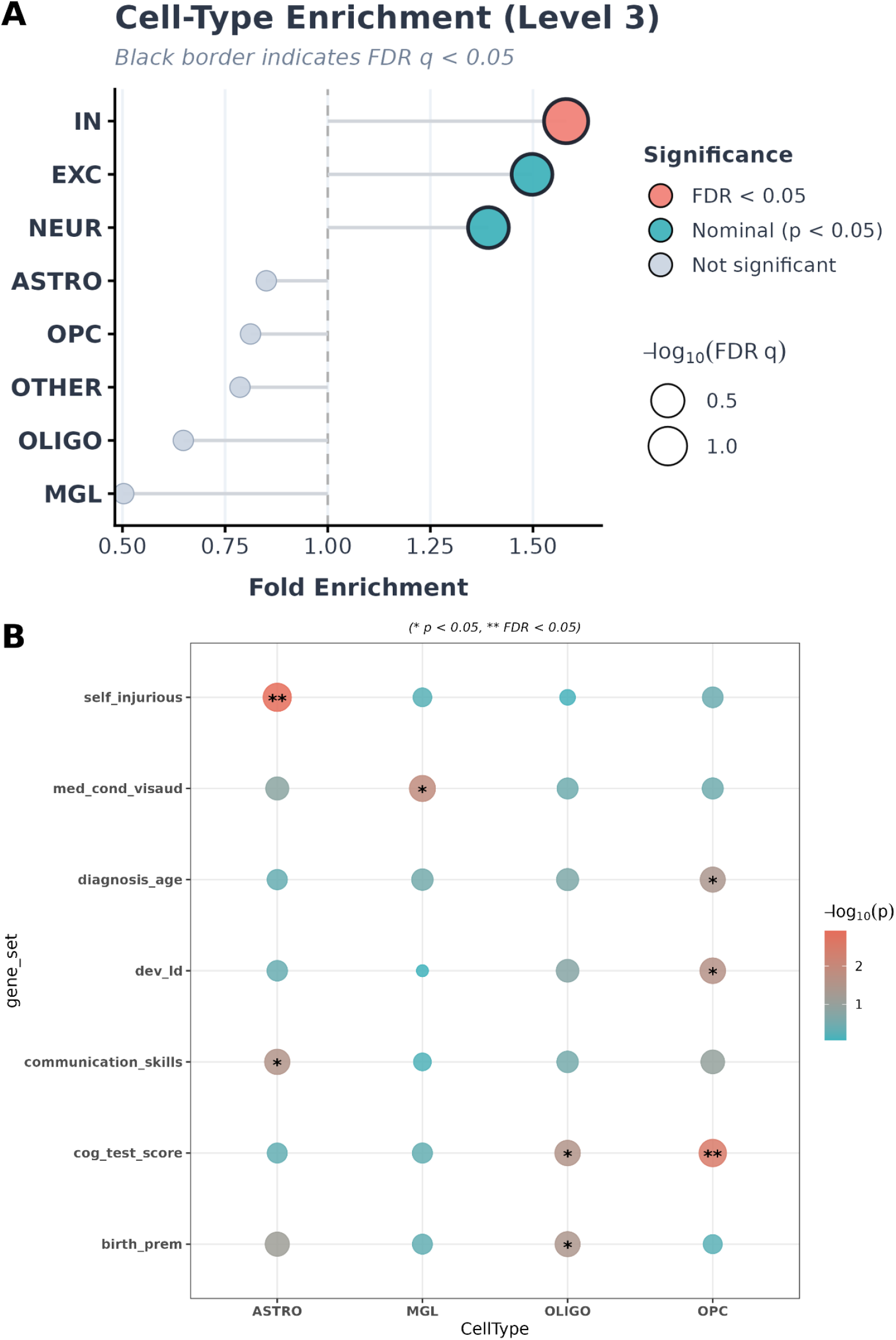
cell type enrichments from EWCE. A: enrichment of the gene sets obtained from the union of GWAS and xWAS genes. B: enrichment of the genes suggestively associated (P < 0.001) with each phenotype. Only cell types and phenotypes with at least one nominal enrichment are shown. In particular, in contrast with the analysis across phenotypes, no neuronal type was enriched in any individual phenotype. IN: inhibitory neurons; EXC: excitatory neurons; NEUR: other neurons (see text and Suppl. Tab. 3); OLIGO: oligodendrocytes; OPC: oligodendrocyte precursors; ASTRO: astrocytes; MGL: microglia; *: nominal P < 0.05; **: FDR < 0.05.

To analyze the results for individual phenotypes we used the same gene lists used for the Gene Ontology analysis (based on nominal P < 0.001). Neurons were not enriched in any phenotype-specific analysis, whereas two glial cell types showed significant enrichment for individual phenotypes (OPCs for cognitive test score, astrocytes for self-injurious behavior; Fig. 4B). Given the permissive threshold used to define these gene sets, these results should be considered exploratory.

At the most granular level only one association survived multiple testing correction, namely that of cluster CGE_291 with communication skills. Several other clusters of caudal ganglionic eminence (CGE) interneurons are also nominally enriched in the genes associated with this factor.

Together, these results suggest genes associated through common variants with phenotypic heterogeneity are preferentially expressed in neurons, particularly interneurons, whereas specific non-neuronal cell types are implicated for individual phenotypes.

#### Developmental stages

Several recent papers suggest that ASD genes can be classified into a few functional programs characterized by different developmental trajectories [59–61]. While based on different approaches ranging from the functional classification of rare causative variants to the transcriptomic analysis of mouse models, these approaches consistently identify a “synaptic” program characterized by higher expression in post-natal than in fetal brain, and a “gene regulation” program characterized by the opposite developmental trend. Other programs identified are more dependent on the specific approach (such as the “morphogenesis” program in [61] or the “signaling” program in [59]).

We asked in which of these programs, if any, the genes associated with phenotypic heterogeneity would fit, using the BrainSpan Atlas of the Developing Human Brain [62]. After grouping the BrainSpan samples into five age groups we applied GSEA with the genes associated with phenotypic heterogeneity by GWAS or xWAS as the gene set, and z-score transformed expression as the ranking score (see Methods). Only genes associated with at least one GRF were considered. This analysis was used to compare the developmental trajectories of our genes with those of the three programs defined in [61].

The genes identified by GWAS (P < 5e-8) or xWAS (FDR < 0.1) displayed a developmental trajectory very similar to that of the “Synapse” program, and clearly distinct from those of the other two programs, in agreement with their functional characterization (Fig. 5A). It should be noted that, since our GRF predictive models are based on adult tissues, genes whose genetic regulation is restricted to fetal development are likely underrepresented among xWAS associations, which could bias the results towards the synaptic trajectory. However, the GWAS genes, which are not subject to this bias, have a qualitatively similar, although not significant, trajectory (Fig. 5A), and the analysis was restricted to genes with at least one GRF, suggesting that such bias did not have a dominant effect on the results. Predictive models trained on fetal brain data [9,10] could address this limitation directly.

**Fig. 5:**
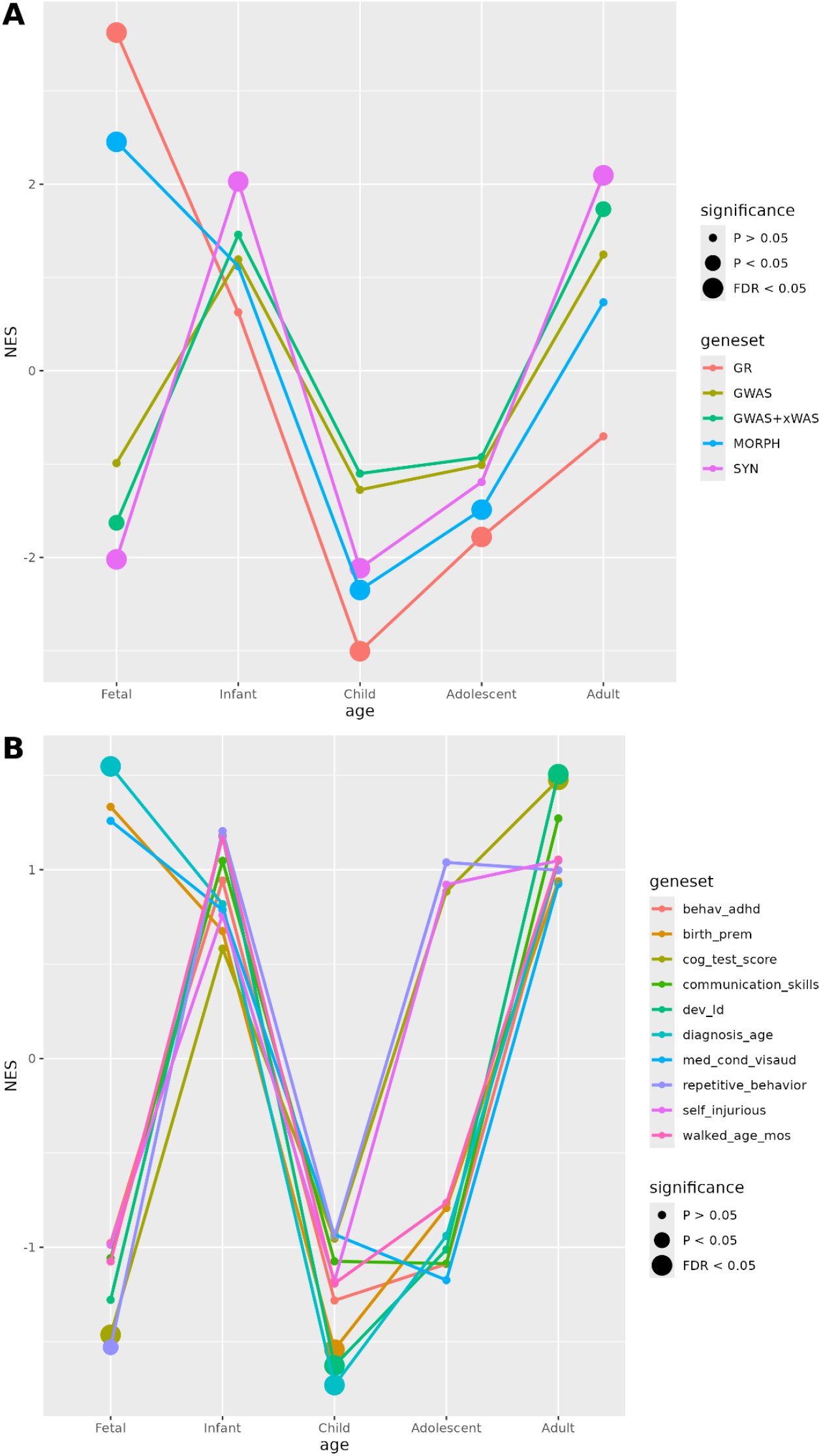
A. Normalized Enrichment Score (NES) trajectories of the GWAS genes and of the union of GWAS and xWAS genes across BrainSpan age groups, compared with those of the “Synapse” (SYN), “Gene Regulation” (GR), and “Morphogenesis” (MORPH) programs of [61]. B. NES trajectories of the genes suggestively associated by xWAS (P < 0.001) with 10 heritable phenotypes. Fetal (8 pcw - 37 pcw); Infant (4 mos - 1 yrs); Child (2 yrs - 11 yrs); Adolescent (13 yrs - 18 yrs); Adult (19 yrs - 40 yrs)

When looking at genes suggestively associated by xWAS (P < 0.001) with each individual phenotype (Fig. 5B), we saw that most phenotypes showed a trajectory similar to the synaptic program, which reached significance for the cognitive phenotypes (cognitive test score and learning disability, FDR < 0.05 at the adult stage).

Conversely, genes associated with age at diagnosis were significantly enriched in the fetal brain (FDR < 0.05), and premature birth and visual/auditory impairment showed a similar but non-significant trend. While premature birth is difficult to interpret due to the lack of distinction between medically induced and spontaneous premature birth in the data, it is natural to expect a relevance of fetally expressed genes to this phenotype. Regarding visual/auditory impairments, a fetal enrichment is compatible with the fact that sensory organs and pathways largely develop prenatally (see e.g. [63,64] for reviews). The heritability of age at ASD diagnosis has been recently dissected in [65], where two different and modestly correlated factors are associated with early and late diagnosis: One could speculate that the former factor could act through fetally expressed genes.

Together, these results suggest that common variants exert their effect on the phenotypic heterogeneity of ASD, and in particular cognitive phenotypes, by acting in particular on genes preferentially expressed in the post-natal brain.

#### Computational drug repositioning

xWAS provides signed, genetically regulated expression signatures for each phenotype. We reasoned that these could be combined with transcriptomic signatures associated with chemical perturbations to identify compounds able to reverse these signatures, and that could hence be considered as potential therapeutic agents. The use of genetically predicted features, rather than measured ones, in this context could in principle present the advantage of removing reverse causality (where the phenotype causes gene expression alterations rather than vice versa). We used the LINCS 2020 (Touchstone v2) dataset [66,67], the latest and most comprehensive release of the Broad Institute’s perturbational gene expression resource, containing over 3 million gene expression profiles, which in particular includes neuronal (NEU) and neural progenitor cell (NPC) models.

Focusing on core ASD dimensions, these were combined with the transcriptional signatures obtained by xWAS for the three heritable ASD factors (repetitive behavior, self-injurious behavior, and communication skills). For each factor we built a signature containing 100 genes positively associated with the factor and 100 negatively associated. Briefly (see Methods) we considered all xWAS data except splicing (for which the sign of the association coefficient is not readily interpretable), and identified for each gene the feature (mRNA expression, enhancer activity, or protein expression) with the largest association z-score (in absolute value) with the factor; associated the corresponding (signed) z-score to the gene; and finally considered the 100 genes with the highest positive (negative) z-score as the “up” and “down” components of the signature.

We then used the “sig_query_l1000” tool provided by LINCS/CLUE to scan for chemical perturbations whose effect on the transcriptome matched or opposed our signatures, limiting the analysis to NEU and NPC cells. The tool outputs, for each signature, a normalized connectivity score (NCS) expressing how strongly, and in which direction, the chemical perturbation matches the query signature. In Fig. 6A we show the compounds with |NCS| > 1 in at least two of the three factors. Compounds with NCS > 1 mimic the query signature, while those with NCS < -1 reverse it, so that the latter can be considered as candidates for therapeutic intervention. None of these connections reached FDR < 0.05; however, a recent study of schizophrenia it was shown [68] that simply filtering for |NCS| > 1 produced candidate compounds that were successfully validated experimentally, irrespective of their FDR.

**Fig. 6:**
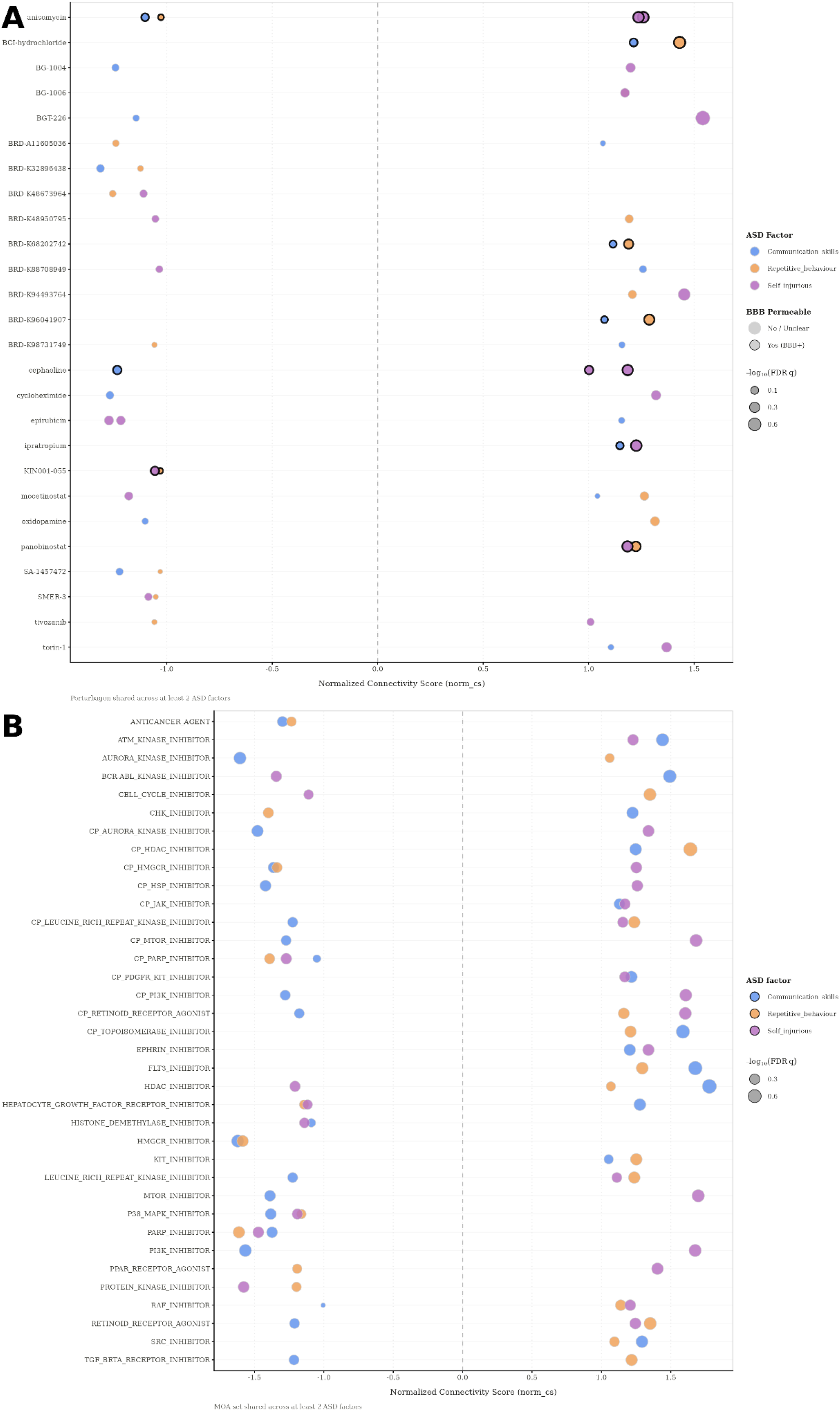
Perturbagen Connectivity Plot: A. Compounds predicted to reverse (NCS < -1) or mimic (NCS > 1) the signature of two or more ASD factors. The bold circles represent compounds able to cross the blood-brain barrier according to established CNS-penetration physicochemical criteria (MW < 500 Da, TPSA < 90 Å² and 1< log P < 5). Compounds can have more than one NCS value (one for neurons and one for neuronal precursor cells). B. Mode-of-action and compound-class level analysis. Classes starting with “CP_” are CMap compound classes, the other ones are MOAs

No single compound was found to reverse the transcriptional signature of all three factors. Three compounds were predicted to reverse both behavioral phenotypes (repetitive and self-injurious behavior), namely a compound of unknown mode of action (BRD-K48673964), a kinase inhibitor (KIN001-055), and SMER-3, an E3 ubiquitin ligase inhibitor. KIN001-055 is predicted to pass the blood-brain barrier (see Methods).

As results on individual compounds are difficult to interpret, we performed the analysis at the compound-class and mode-of-action (MOA) levels, using the same tool (Fig. B). Here we found two classes of compounds predicted to reverse the signatures of all three factors, namely PARP inhibitors and p38 MAPK inhibitors. Compounds of these classes have been recently shown to rescue ASD core phenotypes in mouse models [69,70].

Importantly, some compounds and MOAs show opposite effects on the transcriptional signatures of the three factors. For example the history deacetylase (HDAC) inhibitor mocetinostat, which has been proposed as a therapeutic agent thanks to its ability to normalize *GTF2I* expression in patient-derived cortical neurons of individuals with ASD [71], appears to reverse the self-injurious behavior signature while mimicking those of communication skills and repetitive behavior. The same pattern is shown by HDAC inhibitors as a class in the MOA-level analysis. Also mTOR inhibitors, the most extensively studied pharmacological approach in syndromic ASD [72,73], showed a similarly divergent pattern, reversing the communication skills signature while mimicking the self-injurious behavior signature. It is tempting to speculate that such divergent effects could contribute to the limited efficacy observed in clinical trials despite robust preclinical evidence.

This analysis represents a preliminary in silico screen: Brain penetrance is only predicted computationally, and tolerability or side effects are not considered. More generally, there is a considerable gap between reversing a transcriptional signature in cultured neurons and modifying a behavioural phenotype. With these caveats, the results suggest that combining genetically predicted molecular features with transcriptional response databases offers a tractable route to generating drug repositioning hypotheses in ASD. Notably, some compound classes act in opposing directions across the three phenotypic dimensions. If, as clinical heterogeneity suggests, individuals differ in which dimension predominates, this would argue against a single transcriptional target for ASD and in favour of interventions stratified by symptom profile.

#### Take-home messages, limitations and future perspectives

Our results indicate that combining GWAS and xWAS analyses can nominate candidate molecular mediators of phenotypic heterogeneity in ASD. GWAS and xWAS identified different genes, associated with largely different phenotypes, which nonetheless converged on related functional categories, most notably synaptic function. The recent literature on ASD distinguishes two major functional classes of ASD genes [59–61]: Those involved in gene regulation and chromatin conformation, which tend to be highly expressed during fetal development; and those involved in synaptic transmission, whose expression increases postnatally. Our results suggest that common variants affect phenotypic heterogeneity predominantly through the latter class of genes, particularly for cognitive phenotypes. Age at diagnosis was a notable exception, being associated with genes preferentially expressed in the fetal brain.

Using genetically predicted molecular features as an intermediate step yields signed gene–phenotype effects, which can be used to define transcriptional signatures and compare them with those of large-scale chemical perturbation screens. This provides a first prioritization step for drug repositioning, identifying compounds and compound classes predicted to reverse phenotype-associated transcriptional profiles. Notably, several compound classes, including HDAC and mTOR inhibitors, showed divergent effects across phenotypic dimensions, suggesting that individuals with ASD with different phenotypic profiles might benefit from stratified therapeutic strategies.

A main limitation of our study is the sample size (9,332 genotyped individuals), which likely explains the small number of genome-wide significant discoveries; none of the GWAS loci survive correction for the number of phenotypes tested, and no replication cohort was available. have a distinctive genetic architecture, with a large mutational target and GWAS hits of limited statistical significance [74]. This leads to the expectation that future increases in sample sizes could dramatically increase the number of hits, as has happened e.g. for major depressive disorder in the last decade [74]. In addition, phenotypes were parent-reported and in some cases composite, and the drug repositioning analysis relied on signatures of modest statistical strength and on cell-line perturbation data, so its results should be regarded as hypotheses for experimental testing.

A second limitation concerns the predictive models. Being trained on bulk transcriptomic and proteomic data, they do not directly resolve the cell types involved; while EWCE analysis could be used to indirectly infer the cell types mediating these genetic effects, predictive models directly derived from single-cell analyses are likely to provide more reliable and fine-grained results. Such models, based on large cohorts, have very recently become available [75] and their use represents a promising future direction of research. Moreover, since most models were trained on adult tissues, genes whose genetic regulation is restricted to fetal development are likely underrepresented among xWAS associations. While the consistency of GWAS and xWAS results and our restricted-background analyses suggest that this bias does not dominate our conclusions, models trained on fetal brain data [9,10] could address it directly.

Finally, while our study confirms the relevance of common variants to phenotypic heterogeneity in ASD, other genetic alterations such as rare and copy-number variants are likely to play an important role and could be analyzed in future research within a similar analytical framework, in particular through joint modeling of polygenic scores and rare-variant burden using the whole-exome and whole-genome sequencing data available for the SPARK cohort.

## Conclusions

In summary, we have shown that the combined use of GWAS and xWAS methods based on a variety of intermediate molecular features can help dissect the genetic architecture of phenotypic heterogeneity in ASD. The analysis revealed that a specific functional class of genes, involved in synaptic transmission and expressed predominantly postnatally, mediates a large part of the effects of common variants on such heterogeneity. The xWAS approach also allows screening for compounds able to reverse the transcriptional signatures related to different axes of the phenotypic spectrum, identifying candidates for stratified drug repurposing.

## Materials and Methods

### Subject selection

We conducted analyses using data of individuals with ASD enrolled in the SPARK cohort. The SPARK database gathered participants’ phenotypic data through the completion of detailed surveys on medical, behavioral, and developmental aspects. From these, we selected only the RBS (Repetitive Behaviour Scale) and the SCQ (Social Communication Questionnaire). For factor analysis, we selected all the individuals with a high confidence ASD diagnosis who had completed both phenotypic measures and had not withdrawn from the study, resulting in a sample of 23,939 participants. Among them, 9,332 individuals were retained in the subsequent analyses according to the following criteria: availability of genotype data; reported as white race; and removal of relatives by retaining one individual per family.

### Factor analysis

#### Phenotypes

Factor analysis was conducted using the *Social Communication Questionnaire* SCQ [76] and the *Revised Repetitive Behavioral Scale* RBS [77] questionnaires. The SCQ consists of 40 binary (yes/no) questions mainly focused on measuring social difficulties and, to a lesser extent, repetitive and/or restricted behaviours, as reported by the caregiver. SPARK provides the Lifetime version, which evaluates the individual’s entire history across their lifespan. Specifically, items 1–19 refer to lifetime behaviors, whereas items 20–40 focus on behaviors observed at age 4 or within the past 12 months for younger individuals.

The RBS is also a caregiver-reported questionnaire assessing the presence and severity of restricted and repetitive behaviours, including motor stereotypies, compulsive and ritualistic behaviors, repetitive self-injury, inflexibility and rigidity, as well as circumscribed and intense interests or activities. It comprises 43 questions rated on a four point Likert scale, where higher scores indicate greater severity of autistic traits (0 = behavior does not occur; 1 = mild problem; 2 = moderate problem; 3 = severe problem).

#### Exploratory Factor analysis

Exploratory factor analysis (EFA) [78] helps simplify complex data by grouping correlated variables into underlying factors, facilitating its interpretation. Specifically, it was conducted on 23,939 individuals with ASD having both questionnaires measures, using the R package ‘psych’ [79]. The total number of variables was 82, obtained by merging RBS with SCQ and by removing item 1 of SCQ, which showed no variability.

Before conducting the factor analysis, preliminary statistical evaluations were performed to ensure data suitability and optimal model performance. The first assessment was the Kaiser-Meyer-Olkin (KMO) which measures the sampling adequacy of each observed variable in the model as well as the complete model. KMO is calculated based on the correlation between the variables and it ranges from 0 to 1, with higher values indicating well correlated variables possibly sharing a common underlying factor. In our dataset, for each variable the KMO exceeded the recommended threshold of 0.5; therefore, no variables were excluded. As an additional validation, we performed Bartlett’s test of sphericity to assess whether the correlation matrix significantly differed from an identity matrix. The test yielded a p-value of 0, allowing rejection of the null hypothesis and confirming the data’s suitability for factor analysis.

The optimal number of factors to be extracted was determined by examining the scree plot, performing a parallel analysis and evaluating the interpretability of candidate solutions after applying the *promax* oblique rotation. As a result, we obtained a six-factor model, whose initial fit was assessed with the root mean square error of approximation (RMSEA) and the root mean square of the residual (RMSR), both of which were below the acceptable threshold of 0.08 (0.039 and 0.03, respectively).

#### Confirmatory Factor Analysis

After performing EFA, a confirmatory factor analysis (CFA) [80] was conducted to further evaluate how well the final model fit the data. The fit indices considered included the CFI (Comparative Fit Index), TLI (Tucker-Lewis Index), RMSEA (Root Mean Square Error of Approximation), and SRMR (Standardized Root Mean Square Residual).

The final model solution obtained from EFA yielded six factors loaded to a set of variables with varying strengths. Factor loadings were retained only when higher than 0.4, implying meaningful associations with the latent structure identified. Factor names were assigned based on the underlying concepts of the items that loaded onto them.

CFA fit measures (table X) indicate an overall acceptable fit of the final model. The RMSEA (0.045) and SRMR (0.048) values are both below 0.05 statistical threshold, suggesting a good approximation of the model to the data. The CFI (∼0.8) and TLI (∼0.8) were slightly below the optimal threshold (0.09), but given the large sample size, they still represent a reasonable fit. Notably, the six-factor model described is consistent with the model reported by Warrier et al. [81], providing an additional validation of our results.

### Genotype imputation

The SPARK cohort contains 153,772 individuals aged 1 to >89 years at enrollment with a professional diagnosis of ASD/autism. SPARK provides both whole-exome and whole-genome sequencing data, along with SNP genotyping data, either via Illumina Global Screening Arrays (GSA) or through genotyping-by-sequencing (GxS) [82].

For our analyses we used the SNP genotyping array data from 69,758 subjects. SNPs arrays do not cover the whole genome; instead, they capture common variants across populations leaving millions of variants uncovered. To expand our data, we performed genotype imputation using the TOPMed Reference panel [83], which infers genotypes at untyped SNPs based on LD patterns. In addition, SPARK defines the ‘ref_allele’ as the allele with highest frequency, indicating they may not be based on the actual reference genome. Therefore, prior to imputation, approximately 80.000 alleles were switched, and the corresponding genotype corrected. The initial number of SNPs in our data was 650.206, which increased to 315.167.017 after imputation. The squared correlation (R²) of allele frequency between the imputed samples and the reference panel was 0.964, suggesting nearly perfect performance. A final quality filter was applied to reduce the dataset size, retaining 17.502.29 variants with R² > 0.5 and allele frequency (MAF) > 0.001.

### Phenotype selection

GWAS and xWAS analyses were performed for the scores of the six factors and binary variables expressing the presence of diagnosed co-occurring conditions. These range from neurodevelopmental and psychiatric disorders related to ASD, such as ADHD, intellectual disability, bipolar disorder, cognitive impairment, and language delay, to other medical conditions, including congenital heart disease, renal agenesis, and lung malformations. Specifically, we retained only those diagnoses present in at least 10% of the individuals included in the preceding analyses, resulting in 22 conditions across 9,295 subjects. The retained co-occurring conditions are described in Tab. 4. Moreover, we considered three additional continuous phenotypic variables reported in SPARK, namely age at diagnosis, age at first walking and cognitive score test. For the latter we used the parent-reported IQ range, because it is available for a larger subset of the cohort than the full-scale IQ from clinical records, and has been shown to agree well with standardized measures of IQ range [13]. This score is reported in bins: We transformed it into a continuous variable by taking the midpoint of each bin, except for open ended upper bounds (>130), to which we assigned a value of 135. In total, therefore, we analyzed 31 phenotypes, including 9 continuous and 22 binary ones.

**Tab. 4:** Co-occurring diagnoses occurring in at least 10% of the individuals we study, and used as binary phenotypes in our association analyses.

| Condition ID | Description |
| --- | --- |
| **attn_behav** | Attention and behavior disorder |
| **behav_adhd** | ADHD (Attention Deficit-Hyperactivity Disorder) or ADD |
| **birth_prem** | Premature birth (delivery before 37 weeks) |
| **dev_id** | Intellectual disability, cognitive impairment, global developmental delay, or borderline intellectual functioning |
| **dev_lang** | Speech and language, intellectual disability/cognitive Impairment, learning disability (LD) or other developmental delay or developmental disability |
| **dev_lang_dis** | Language delay or language disorder |
| **dev_ld** | Learning disability (LD, learning disorder, including reading, written expression, math, or NVLD (Nonverbal learning disability)) |
| **dev_motor** | Motor delay (e.g., delay in walking) or developmental coordination disorder |
| **dev_soc_prag** | Social (Pragmatic) Communication Disorder |
| **dev_speech** | Speech articulation problems |
| **eating_probs** | Problems with eating foods - not diagnosed by a professional |
| **feeding_dx** | Feeding/eating problems |
| **med_cond_birth** | Birth or pregnancy complications |
| **med_cond_growth** | Growth conditions (including height, weight and head size) |
| **med_cond_neuro** | Neurological conditions |
| **med_cond_visaud** | Visual or auditory impairments. |
| **mood_anx** | Anxiety disorder, such as panic, phobia, agoraphobia, or generalized anxiety disorder (GAD) except for social anxiety |
| **mood_or_anx** | Mood, Depression, Anxiety or OCD |
| **mood_soc_anx** | Social Anxiety Disorder/Social Phobia |
| **sleep_dx** | Sleep Disorder or sleep problem diagnosed by a professional |
| **sleep_eat_toilet** | Sleep, Feeding/Eating or toileting problem/s |
| **sleep_probs** | Sleep problems not diagnosed by a professional |

### Genome-wide association studies

For the association we used REGENIE [84]: REGENIE performs GWAS chromosome-by-chromosome including polygenic predictions from other chromosomes for robust correction of population stratification. Variants were filtered prior to polygenic model generation to retain only well-imputed and high-quality markers. We restricted the set of variants with imputation INFO scores > 0.90 and applied linkage disequilibrium (LD) pruning (2000 Kb window with 1 SNPs as spacing, r^2^ > 0.5) to obtain a subset of variants in approximate LD equilibrium. This reduced the set of variants for REGENIE step 1 to 781,093 SNPs. The polygenic predictors estimated in step 1 were then used in REGENIE step 2 to perform GWAS on all variants with MAF > 0.01. Sex, age at enrollment and the first 10 genomic PCs were used as covariates. FUMA [14] was used to map variants to genes.

LDSC [23] was used on the summary statistics of the GWAS studies to estimate the SNP heritability of each trait. For the xWAS analysis we chose the 10 traits showing at least some evidence of heritability (z(h^2^) > 1.5).

### Predictive models of genetically regulated molecular features

The molecular phenotypes of our interest include gene and protein expression, splicing and enhancer RNAs.

#### Gene expression

We used the Joint-Tissue Imputation (JTI) models [30] for 15 relevant tissues, 13 of which were brain regions: Amygdala, Anterior Cingulate Cortex (BA24), Caudate (basal ganglia), Cerebellar Hemisphere, Cerebellum, Cortex, Frontal Cortex (BA9), Hippocampus, Hypothalamus, Nucleus Accumbens (basal ganglia), Putamen (basal ganglia), Spinal Cord (cervical C1), and Substantia Nigra. The remaining tissues were EBV-transformed lymphocytes and Whole Blood, given the evidence of immune dysregulation in autism [85]. Alongside these models, we also included the JTI PsychENCODE [86] model for the prefrontal cortex. The choice of JTI relies on its novel approach based on leveraging shared genetic regulation across different tissues, to improve prediction performance in a tissue-dependent manner [87].

#### Splicing

We used PrediXcan elastic net splicing prediction models [88] for the same tissues described in the previous section, with the exception of the PsychENCODE prefrontal cortex model, which is not available.

#### Protein expression

We used the FUSION models produced by Wingo et al. [89] from a multi-ancestry brain pQTL fine-mapping of 1,362 donors, covering a total of 3,771 genes. All predictive models (top1, blup, lasso, enet and bslmm) were used independently [90].

Additionally, we retrieved from the OmicsPred resource [53] polygenic scores of plasma protein abundance. In particular, we selected genetic scores for 2,612 Olink UK Biobank proteins derived from individuals of European ancestry, as well as scores for 2,384 proteins measured using the SomaScan platform.

#### Enhancer activity

Betti et al. [92] developed JTI models of genetically regulated expression of enhancer RNAs across 49 cell and tissue types using data from the Human enhancer RNA Atlas (HeRA, https://hanlab.uth.edu/HeRA/) [93]. For our analyses, we used the models corresponding to the same 13 brain tissues, whole blood, and lymphocytes described above; these models were retrieved from Zenodo under accession code 14027849 [94]. Enhancer/gene associations were derived separately for each tissue by selecting the gene with the highest correlation with the enhancer in each tissue, based on data available from the HeRA atlas.

### Prediction of the genetically regulated molecular phenotypes

The prediction of the intermediate molecular phenotypes described above was performed across all tissues for the 9,332 selected individuals using the predict.py tool obtained from the MetaXcan GitHub repository (https://github.com/hakyimlab/MetaXcan.git). We discarded all features for which the number of SNPs in the genotype data was less than 80% of those included in the prediction model.

### Association between genetically predicted molecular traits and phenotypes

The association between the predicted molecular phenotypes and the phenotypes was assessed using the *‘lm’* or *‘glm’* function in R for continuous and binary phenotypes, respectively. For each predictive model, the phenotype value was regressed against the predicted value of each GRF, including age, sex and 10 genomic PCs as covariates. Association P-values were corrected with the Benjamini-Hochberg procedure, separately for each trait and type of GRF (gene expression, protein expression, splicing, and eRNA) but considering together all tissues (for brain proteomics: all FUSION predictive models; for plasma proteomics: both platforms).

GRFs significantly associated (FDR < 0.1) with phenotypes were organized into approximately independent clusters, that we call “loci”, by computing all their pairwise correlations to build a graph in which two GRFare joined by an edge if their squared correlation is > 0.5. The approximately independent loci were defined as the connected components of thai graph. The same procedure was applied to the GRFs with suggestive evidence of association (P < 0.001) used in the phenotype-specific enrichment analyses.

### Enrichment analyses

Enrichment analyses require gene sets, while the loci identified by both GWAS and xWAS often contain several candidate genes. To assign a unique gene to each locus in a data-driven way, allowing unbiased enrichment analyses, we proceed as follows. For each GWAS locus we associated the gene associated to it by FUMA, if unique; otherwise, we chose the nearest gene, if unique; no gene was associated to loci for which neither the FUMA gene nor the nearest gene were unique. To each xWAS locus we associated the gene linked to the lead GRF, i.e. the GRF with the most significant association P-value.

The gene sets used across the enrichment analyses described below were: (a) all genes associated with genome-wide significant GWAS loci; (b) all genes associated by xWAS analysis to a phenotype with FDR < 0.1; (c) the union of (a) and (b); (d) for each heritable phenotype, all genes associated with GRFs with nominal association P-value < 0.001.

#### Gene Ontology

Gene Ontology enrichment analysis was performed with the enrichGO function of the clusterProfiler package [95], which implements exact Fisher overrepresentation test with Benjamini-Hochberg correction for multiple testing. Enriched terms were not considered if the enrichment derived from a single gene.

#### BrainSpan

We retrieved from [BrainSpan](https://www.brainspan.org/) the transcriptome of 26 brain structures across the full course of human brain development, from 8 pcw to 40 years old. Gene expression data were first averaged across all samples within predefined developmental stages: Fetal (8 PCW–37 PCW), Infant (4 months–1 year), Child (2–11 years), Adolescent (13–18 years), and Adult (19–40 years). We limited the analysis to genes associated with at least one GRF in our xWAS study. Expression values for each gene were transformed into z scores, which were used to perform Gene Set Enrichment Analysis with the GSEA function of the clusterProfiler package [95], using our gene sets against the gene lists ranked by expression z score. Normalized Enrichment Scores (NES) were used to plot the developmental trajectories.

#### Expression Weighted Cell-type Enrichment

Expression Weighted Cell-type Enrichment (EWCE) [96] is a statistical technique that determines whether a list of genes is overexpressed in a specific cell type, compared to a background distribution. Here, the tested gene sets were the ones used for all enrichment analyses, while the background set was derived from the Human Brain Cell Atlas [97]., which is organized into 30 superclusters, 461 clusters, and 3313 subclusters. We manually created an additional annotation level consisting of 8 “megaclusters” based, for neurons, on the predominant neurotransmitter expressed by the original cluster. We thus defined the following megaclusters: oligodendrocyte precursor cells (OPCs), oligodendrocytes (OLIGO), astrocytes (ASTRO), microglia (MGL), inhibitory neurons (IN), excitatory neurons (EXC), neurons (NEUR), and other (OTHER). The NEUR megacluster includes clusters expressing a mix or neuromodulatory neurotransmitter profiles, including glycine, VGLUT3, serotonin, and acetylcholine. The OTHER category contains a mix of cell types, primarily ependymal and vascular cells. The assignment of each cluster to a megacluster is shown in Suppl. Tab. 3. The background cell-type dataset used for the analysis included a gene-by-cell expression matrix along with three levels of cell-type annotation: 461 clusters, 31 superclusters, and 8 megaclusters. Enrichment analysis for each annotation level was finally performed in R using the ‘EWCE’ Bioconductor package [98].

### Drug repurposing

To identify potential therapeutic candidates targeting the heritable core factor signatures (Self-Injurious Behavior, Repetitive Behavior, and Communication Skills), we executed a local Connectivity Map ([CMap](https://www.broadinstitute.org/connectivity-map-cmap)) [66] pipeline using the LINCS2020 dataset, the latest and most comprehensive release of the Broad Institute’s perturbational gene expression resource, containing over 3 million gene expression profiles. Unlike the earlier LINCS Phase 1 release, the LINCS 2020 dataset includes neuronal (NEU) and neural progenitor cell (NPC) models, providing substantially greater biological relevance for neuropsychiatric applications.

The transcriptional input signatures for each factor were built from the xWAS results by selecting the top 100 positively associated and top 100 negatively associated genes ranked by t-value. We excluded the splicing signatures since the sign of the association coefficient is not readily interpretable. The analysis aims to assess the degree to which the up-regulated query genes (q-up) appear toward the top of the rank-ordered signature and the down-regulated query genes (q-down) appear toward the bottom of the perturbational signature (positive connectivity) or vice-versa (negative connectivity). The metric used to interpret the result is called Weighted Connectivity score (WTCS), a composite, bi-directional version of the weighted Kolmogorov-Smirnov enrichment statistic (ES). WTCS was subsequently normalized within each cell line and perturbagen type to produce a normalized connectivity score (NCS). The result compounds list was finally filtered retaining only chemical compounds with |NCS| > 1 and Transcriptional Activity Score (TAS) *> 0.4,* which reflects the overall magnitude and reproducibility of a compound’s biological effect. While positive *NCS* values designate compounds that mimic the factor signature, negative *NCS* values identify signature-inverting compounds as candidate therapeutic targets.

Moreover, we evaluated the potential of candidate compounds to cross the blood-brain barrier (*BBB*) based on established CNS-penetration physicochemical criteria: molecular weight (MW < 500 Da), topological polar surface area (TPSA < 90 Å²), and lipophilicity (log P between 1 and 5). These thresholds favor passive transcellular diffusion by balancing molecular size, restricting hydrogen-bonding desolvation penalties, and optimizing lipid-bilayer partitioning.

In addition to individual compounds evaluations, we conducted a Mechanism of Action (MoA)-level connectivity analysis using the provided mean consensus signature profiles derived from compounds that share the same MoA. This analysis followed the identical analytical pipeline, normalization scheme, and interpretation criteria described above.

## Data Availability

All data produced in the present work are contained in the manuscript and in the Supplementary Material

## Acknowledgements

SPARK cohort data (phenotypic and genotyping) were accessed under project no. 1517.1. Camilla Marinelli is a PhD student enrolled in the National PhD in Artificial Intelligence (XL cycle) course on Health and life sciences, organized by Università Campus Bio-Medico di Roma.

## Supplementary Information

**Suppl. Tab. 1:** xWAS associations at FDR < 0.1

**Suppl. Tab. 2:** GO enrichment analysis.

**Suppl. Tab. 3**: Three annotation levels from Human Brain Atlas used for EWCEanalysis. In particular, the ‘megacluster’ represents the third-level, custom annotation and includes eight major cell types defined according to their neurotransmitter expression.

## Notes

### Competing Interest Statement

The authors have declared no competing interest.

### Author Declarations

The University of Turin Bioethics Committee gave ethical approval for this work on October 15 2024

## References

1. Lord C, Brugha TS, Charman T, Cusack J, Dumas G, Frazier T, et al. Autism spectrum disorder. Nat Rev Dis Primer. 2020;6: 5. doi:10.1038/s41572-019-0138-4

2. Havdahl A, Niarchou M, Starnawska A, Uddin M, van der Merwe C, Warrier V. Genetic contributions to autism spectrum disorder. Psychol Med. 2021;51: 2260–2273. doi:10.1017/S0033291721000192

3. Warrier V, Zhang X, Reed P, Havdahl A, Moore TM, Cliquet F, et al. Genetic correlates of phenotypic heterogeneity in autism. Nat Genet. 2022. doi:10.1038/s41588-022-01072-5

4. Litman A, Sauerwald N, Green Snyder L, Foss-Feig J, Park CY, Hao Y, et al. Decomposition of phenotypic heterogeneity in autism reveals underlying genetic programs. Nat Genet. 2025;57: 1611–1619. doi:10.1038/s41588-025-02224-z

5. SPARK Consortium. Electronic address, SPARK Consortium. SPARK: A US Cohort of 50,000 Families to Accelerate Autism Research. Neuron. 2018;97: 488–493. doi:10.1016/j.neuron.2018.01.015

6. Lee I-H, Koelliker E, Kong SW. Quantitative trait locus analysis for endophenotypes reveals genetic substrates of core symptom domains and neurocognitive function in autism spectrum disorder. Transl Psychiatry. 2022;12: 407. doi:10.1038/s41398-022-02179-3

7. Huang K, Wu Y, Shin J, Zheng Y, Siahpirani AF, Lin Y, et al. Transcriptome-wide transmission disequilibrium analysis identifies novel risk genes for autism spectrum disorder. PLoS Genet. 2021;17: e1009309. doi:10.1371/journal.pgen.1009309

8. Gandal MJ, Zhang P, Hadjimichael E, Walker RL, Chen C, Liu S, et al. Transcriptome-wide isoform-level dysregulation in ASD, schizophrenia, and bipolar disorder. Science. 2018;362: eaat8127. doi:10.1126/science.aat8127

9. Wen C, Margolis M, Dai R, Zhang P, Przytycki PF, Vo DD, et al. Cross-ancestry, cell-type-informed atlas of gene, isoform, and splicing regulation in the developing human brain. medRxiv: The Preprint Server for Health Sciences; 2023. doi:10.1101/2023.03.03.23286706

10. Walker RL, Ramaswami G, Hartl C, Mancuso N, Gandal MJ, de la Torre-Ubieta L, et al. Genetic Control of Expression and Splicing in Developing Human Brain Informs Disease Mechanisms. Cell. 2019;179: 750–771.e22. doi:10.1016/j.cell.2019.09.021

11. Lam KSL, Aman MG. The Repetitive Behavior Scale-Revised: independent validation in individuals with autism spectrum disorders. J Autism Dev Disord. 2007;37: 855–866. doi:10.1007/s10803-006-0213-z

12. Rutter M, Bailey A, Lord C. The Social Communication Questionnaire: Manual. Los Angeles: Western Psychological Services; 2003.

13. Lee CM, Green Snyder L, Carpenter LA, Harris J, Kanne S, Taylor CM, et al. Agreement of parent-reported cognitive level with standardized measures among children with autism spectrum disorder. Autism Res Off J Int Soc Autism Res. 2023;16: 1210–1224. doi:10.1002/aur.2934

14. Watanabe K, Taskesen E, van Bochoven A, Posthuma D. Functional mapping and annotation of genetic associations with FUMA. Nat Commun. 2017;8: 1826. doi:10.1038/s41467-017-01261-5

15. Griswold AJ, Ma D, Sacharow SJ, Robinson JL, Jaworski JM, Wright HH, et al. A de novo 1.5 Mb microdeletion on chromosome 14q23.2-23.3 in a patient with autism and spherocytosis. Autism Res Off J Int Soc Autism Res. 2011;4: 221–227. doi:10.1002/aur.186

16. Pollak RM, Murphy MM, Epstein MP, Zwick ME, Klaiman C, Saulnier CA, et al. Neuropsychiatric phenotypes and a distinct constellation of ASD features in 3q29 deletion syndrome: results from the 3q29 registry. Mol Autism. 2019;10: 30. doi:10.1186/s13229-019-0281-5

17. Dason JS, Romero-Pozuelo J, Atwood HL, Ferrús A. Multiple roles for frequenin/NCS-1 in synaptic function and development. Mol Neurobiol. 2012;45: 388–402. doi:10.1007/s12035-012-8250-4

18. Piton A, Michaud JL, Peng H, Aradhya S, Gauthier J, Mottron L, et al. Mutations in the calcium-related gene IL1RAPL1 are associated with autism. Hum Mol Genet. 2008;17: 3965–3974. doi:10.1093/hmg/ddn300

19. de Rezende VB, Rosa DV, Comim CM, Magno LAV, Rodrigues ALS, Vidigal P, et al. NCS-1 deficiency causes anxiety and depressive-like behavior with impaired non-aversive memory in mice. Physiol Behav. 2014;130: 91–98. doi:10.1016/j.physbeh.2014.03.005

20. Nguyen LD, Nolte LG, Tan WJT, Giuvelis D, Santos-Sacchi J, Bilsky E, et al. Comprehensive somatosensory and neurological phenotyping of NCS1 knockout mice. Sci Rep. 2021;11: 2372. doi:10.1038/s41598-021-81650-5

21. Li J, Xie Y, Cornelius S, Jiang X, Sando R, Kordon SP, et al. Alternative splicing controls teneurin-latrophilin interaction and synapse specificity by a shape-shifting mechanism. Nat Commun. 2020;11: 2140. doi:10.1038/s41467-020-16029-7

22. Saravanapandian V, Nadkarni D, Hsu S-H, Hussain SA, Maski K, Golshani P, et al. Abnormal sleep physiology in children with 15q11.2-13.1 duplication (Dup15q) syndrome. Mol Autism. 2021;12: 54. doi:10.1186/s13229-021-00460-8

23. Bulik-Sullivan BK, Loh P-R, Finucane HK, Ripke S, Yang J, Schizophrenia Working Group of the Psychiatric Genomics Consortium, et al. LD Score regression distinguishes confounding from polygenicity in genome-wide association studies. Nat Genet. 2015;47: 291–295. doi:10.1038/ng.3211

24. GTEx Consortium. The GTEx Consortium atlas of genetic regulatory effects across human tissues. Science. 2020;369: 1318–1330. doi:10.1126/science.aaz1776

25. Wang D, Liu S, Warrell J, Won H, Shi X, Navarro FCP, et al. Comprehensive functional genomic resource and integrative model for the human brain. Science. 2018;362. doi:10.1126/science.aat8464

26. Zhang Z, Hong W, Ruan H, Jing Y, Li S, Liu Y, et al. HeRA: an atlas of enhancer RNAs across human tissues. Nucleic Acids Res. 2021;49: D932–D938. doi:10.1093/nar/gkaa940

27. Wingo AP, Liu Y, Vattathil SM, Gerasimov ES, Mei Z, Ravindran SP, et al. Multiancestry brain pQTL fine-mapping and integration with genome-wide association studies of 21 neurologic and psychiatric conditions. Nat Genet. 2025;57: 2156–2165. doi:10.1038/s41588-025-02291-2

28. Moore C, Sambrook J, Walker M, Tolkien Z, Kaptoge S, Allen D, et al. The INTERVAL trial to determine whether intervals between blood donations can be safely and acceptably decreased to optimise blood supply: study protocol for a randomised controlled trial. Trials. 2014;15: 363. doi:10.1186/1745-6215-15-363

29. Sun BB, Chiou J, Traylor M, Benner C, Hsu Y-H, Richardson TG, et al. Plasma proteomic associations with genetics and health in the UK Biobank. Nature. 2023. doi:10.1038/s41586-023-06592-6

30. Zhou D, Jiang Y, Zhong X, Cox NJ, Liu C, Gamazon ER. A unified framework for joint-tissue transcriptome-wide association and Mendelian randomization analysis. Nat Genet. 2020;52: 1239–1246. doi:10.1038/s41588-020-0706-2

31. Betti MJ, Lin P, Aldrich MC, Gamazon ER. Genetically regulated eRNA expression predicts chromatin contact frequency and reveals genetic mechanisms at GWAS loci. Nat Commun. 2025;16: 3193. doi:10.1038/s41467-025-58023-x

32. Barbeira AN, Dickinson SP, Bonazzola R, Zheng J, Wheeler HE, Torres JM, et al. Exploring the phenotypic consequences of tissue specific gene expression variation inferred from GWAS summary statistics. Nat Commun. 2018;9: 1825. doi:10.1038/s41467-018-03621-1

33. Barbeira AN, Bonazzola R, Gamazon ER, Liang Y, Park Y, Kim-Hellmuth S, et al. Exploiting the GTEx resources to decipher the mechanisms at GWAS loci. Genome Biol. 2021;22: 49. doi:10.1186/s13059-020-02252-4

34. Gamazon ER, Wheeler HE, Shah KP, Mozaffari SV, Aquino-Michaels K, Carroll RJ, et al. A gene-based association method for mapping traits using reference transcriptome data. Nat Genet. 2015;47: 1091–1098. doi:10.1038/ng.3367

35. Gusev A, Ko A, Shi H, Bhatia G, Chung W, Penninx BWJH, et al. Integrative approaches for large-scale transcriptome-wide association studies. Nat Genet. 2016;48: 245–252. doi:10.1038/ng.3506

36. Loesch D. Genetic prediction of UK Biobank proteomics data for the OmicsPred portal. Eur J Hum Genet. 2024;32: 799–903. doi:10.1038/s41431-024-01732-6

37. Xu Y, Ritchie SC, Liang Y, Timmers PRHJ, Pietzner M, Lannelongue L, et al. An atlas of genetic scores to predict multi-omic traits. Nature. 2023;616: 123–131. doi:10.1038/s41586-023-05844-9

38. Yao Y, Hong S, Ikeda T, Mori H, MacDougald OA, Nada S, et al. Amino acids enhance polyubiquitination of Rheb and its binding to mTORC1 by blocking lysosomal ATXN3 deubiquitinase activity. Mol Cell. 2020;80: 437–451.e6. doi:10.1016/j.molcel.2020.10.004

39. Magdalon J, Sánchez-Sánchez SM, Griesi-Oliveira K, Sertié AL. Dysfunctional mTORC1 Signaling: A Convergent Mechanism between Syndromic and Nonsyndromic Forms of Autism Spectrum Disorder? Int J Mol Sci. 2017;18: 659. doi:10.3390/ijms18030659

40. Evert BO, Araujo J, Vieira-Saecker AM, de Vos RAI, Harendza S, Klockgether T, et al. Ataxin-3 represses transcription via chromatin binding, interaction with histone deacetylase 3, and histone deacetylation. J Neurosci Off J Soc Neurosci. 2006;26: 11474–11486. doi:10.1523/JNEUROSCI.2053-06.2006

41. Tseng C-EJ, McDougle CJ, Hooker JM, Zürcher NR. Epigenetics of Autism Spectrum Disorder: Histone Deacetylases. Biol Psychiatry. 2022;91: 922–933. doi:10.1016/j.biopsych.2021.11.021

42. Khaliulin I, Hamoudi W, Amal H. The multifaceted role of mitochondria in autism spectrum disorder. Mol Psychiatry. 2025;30: 629–650. doi:10.1038/s41380-024-02725-z

43. Cheon S, Dean M, Chahrour M. The ubiquitin proteasome pathway in neuropsychiatric disorders. Neurobiol Learn Mem. 2019;165: 106791. doi:10.1016/j.nlm.2018.01.012

44. Arenella M, Cadby G, De Witte W, Jones RM, Whitehouse AJ, Moses EK, et al. Potential role for immune-related genes in autism spectrum disorders: Evidence from genome-wide association meta-analysis of autistic traits. Autism Int J Res Pract. 2022;26: 361–372. doi:10.1177/13623613211019547

45. Pak DT, Yang S, Rudolph-Correia S, Kim E, Sheng M. Regulation of dendritic spine morphology by SPAR, a PSD-95-associated RapGAP. Neuron. 2001;31: 289–303. doi:10.1016/s0896-6273(01)00355-5

46. Matsuura K, Kobayashi S, Konno K, Yamasaki M, Horiuchi T, Senda T, et al. SIPA1L1/SPAR1 Interacts with the Neurabin Family of Proteins and is Involved in GPCR Signaling. J Neurosci Off J Soc Neurosci. 2022;42: 2448–2473. doi:10.1523/JNEUROSCI.0569-21.2022

47. Annamneedi A, Gora C, Dudas A, Leray X, Bozon V, Crépieux P, et al. Towards the convergent therapeutic potential of G protein-coupled receptors in autism spectrum disorders. Br J Pharmacol. 2025;182: 3044–3067. doi:10.1111/bph.16216

48. Emerick MC, Stein R, Kunze R, McNulty MM, Regan MR, Hanck DA, et al. Profiling the array of Ca(v)3.1 variants from the human T-type calcium channel gene CACNA1G: alternative structures, developmental expression, and biophysical variations. Proteins. 2006;64: 320–342. doi:10.1002/prot.20877

49. Chemin J, Siquier-Pernet K, Nicouleau M, Barcia G, Ahmad A, Medina-Cano D, et al. De novo mutation screening in childhood-onset cerebellar atrophy identifies gain-of-function mutations in the CACNA1G calcium channel gene. Brain J Neurol. 2018;141: 1998–2013. doi:10.1093/brain/awy145

50. Skvortsova L, Perfilyeva A, Bespalova K, Kuzovleva Y, Kabysheva N, Khamdiyeva O. 7p22.3 microdeletion: a case study of a patient with congenital heart defect, neurodevelopmental delay and epilepsy. Orphanet J Rare Dis. 2024;19: 301. doi:10.1186/s13023-024-03321-8

51. Genovese A, Cox DM, Butler MG. Partial Deletion of Chromosome 1p31.1 Including only the Neuronal Growth Regulator 1 Gene in Two Siblings. J Pediatr Genet. 2015;4: 23–28. doi:10.1055/s-0035-1554977

52. Tassano E, Gamucci A, Celle ME, Ronchetto P, Cuoco C, Gimelli G. Clinical and Molecular Cytogenetic Characterization of a de novo Interstitial 1p31.1p31.3 Deletion in a Boy with Moderate Intellectual Disability and Severe Language Impairment. Cytogenet Genome Res. 2015;146: 39–43. doi:10.1159/000431391

53. Su F, Pfundstein G, Sah S, Zhang S, Keable R, Hagan DW, et al. Neuronal growth regulator 1 (NEGR1) promotes the synaptic targeting of glutamic acid decarboxylase 65 (GAD65). J Neurochem. 2025;169: e16279. doi:10.1111/jnc.16279

54. Szczurkowska J, Pischedda F, Pinto B, Managò F, Haas CA, Summa M, et al. NEGR1 and FGFR2 cooperatively regulate cortical development and core behaviours related to autism disorders in mice. Brain J Neurol. 2018;141: 2772–2794. doi:10.1093/brain/awy190

55. Zhang Y-Q, Zhang Q, Yang Y, Yu L-L, Fan N-L, Wu Y, et al. Elevated NEGR1 in brain induces anxiety or depression-like phenotypes and synaptic dysfunction. Mol Psychiatry. 2025;30: 4627–4640. doi:10.1038/s41380-025-03052-7

56. Nakamura K, Kubo A, Sanaka S, Kamiya S, Itagaki K, Sasaki T. Cytoskeletal Dynamics and Molecular Motor Dysfunction in Psychiatric Disorders: Insights from Schizophrenia and Autism Spectrum Disorder. Biology. 2026;15: 550. doi:10.3390/biology15070550

57. Skene NG, Grant SGN. Identification of Vulnerable Cell Types in Major Brain Disorders Using Single Cell Transcriptomes and Expression Weighted Cell Type Enrichment. Front Neurosci. 2016;10: 16. doi:10.3389/fnins.2016.00016

58. Siletti K, Hodge R, Mossi Albiach A, Lee KW, Ding S-L, Hu L, et al. Transcriptomic diversity of cell types across the adult human brain. Science. 2023;382: eadd7046. doi:10.1126/science.add7046

59. Roh JD, Jun Y, Jeon H, Kim J, Yi Y, Kim M, et al. Transcriptome-based classification in mice with ASD-risk mutations. Science. 2026;393: 1250–1257. doi:10.1126/science.adz6688

60. Satterstrom FK, Kosmicki JA, Wang J, Breen MS, De Rubeis S, An J-Y, et al. Large-Scale Exome Sequencing Study Implicates Both Developmental and Functional Changes in the Neurobiology of Autism. Cell. 2020;180: 568–584.e23. doi:10.1016/j.cell.2019.12.036

61. Smith RL, Liao Y, Klei L, Zhang L, Ma Y, Tang M, et al. Autism genes converge on three functional programs organized by neuronal subclass, developmental timing, and cortical patterning. bioRxiv; 2026. p. 2026.09.22.753508. doi:10.64898/2026.09.22.753508

62. Li M, Santpere G, Imamura Kawasawa Y, Evgrafov OV, Gulden FO, Pochareddy S, et al. Integrative functional genomic analysis of human brain development and neuropsychiatric risks. Science. 2018;362: eaat7615. doi:10.1126/science.aat7615

63. Quinn PMJ, Wijnholds J. Retinogenesis of the Human Fetal Retina: An Apical Polarity Perspective. Genes. 2019;10: 987. doi:10.3390/genes10120987

64. Moore JK, Linthicum FH. The human auditory system: a timeline of development. Int J Audiol. 2007;46: 460–478. doi:10.1080/14992020701383019

65. Zhang X, Grove J, Gu Y, Buus CK, Nielsen LK, Neufeld SAS, et al. Polygenic and developmental profiles of autism differ by age at diagnosis. Nature. 2025. doi:10.1038/s41586-025-09542-6

66. Subramanian A, Narayan R, Corsello SM, Peck DD, Natoli TE, Lu X, et al. A Next Generation Connectivity Map: L1000 Platform and the First 1,000,000 Profiles. Cell. 2017;171: 1437–1452.e17. doi:10.1016/j.cell.2017.10.049

67. Xie Z, Kropiwnicki E, Wojciechowicz ML, Jagodnik KM, Shu I, Bailey A, et al. Getting Started with LINCS Datasets and Tools. Curr Protoc. 2022;2: e487. doi:10.1002/cpz1.487

68. Deans MPJ, Retallick-Townsley KG, Li A, Seah C, Johnson J, Garcia Gonzalez J, et al. Functional implications of polygenic risk for schizophrenia in human neurons. Nat Commun. 2026. doi:10.1038/s41467-025-67959-z

69. Ahmad SF, Ansari MA, Nadeem A, Bakheet SA, Alqahtani F, Alhoshani AR, et al. 5-aminoisoquinolinone attenuates social behavior deficits and immune abnormalities in the BTBR T+ Itpr3tf/J mouse model for autism. Pharmacol Biochem Behav. 2020;189: 172859. doi:10.1016/j.pbb.2020.172859

70. Robson MJ, Quinlan MA, Margolis KG, Gajewski-Kurdziel PA, Veenstra-VanderWeele J, Gershon MD, et al. p38α MAPK signaling drives pharmacologically reversible brain and gastrointestinal phenotypes in the SERT Ala56 mouse. Proc Natl Acad Sci U S A. 2018;115: E10245–E10254. doi:10.1073/pnas.1809137115

71. Cavallo F, Troglio F, Fagà G, Fancelli D, Shyti R, Trattaro S, et al. High-throughput screening identifies histone deacetylase inhibitors that modulate GTF2I expression in 7q11.23 microduplication autism spectrum disorder patient-derived cortical neurons. Mol Autism. 2020;11: 88. doi:10.1186/s13229-020-00387-6

72. Thomas SD, Jha NK, Ojha S, Sadek B. mTOR Signaling Disruption and Its Association with the Development of Autism Spectrum Disorder. Molecules. 2023;28: 1889. doi:10.3390/molecules28041889

73. Winden KD, Ebrahimi-Fakhari D, Sahin M. Abnormal mTOR Activation in Autism. Annu Rev Neurosci. 2018;41: 1–23. doi:10.1146/annurev-neuro-080317-061747

74. Zhu H, Simons YB, Spence JP, Sella G, Pritchard JK. Genetic architectures of brain-related traits are shaped by strong selective constraints. Proc Natl Acad Sci U S A. 2026;123: e2609814123. doi:10.1073/pnas.2609814123

75. Venkatesh S, Kosoy R, Wu Z, Anyfantakis M, Dillard C, N M P, et al. Single-nucleus transcriptome-wide association study of human brain disorders. Nature. 2026;657: 1016–1026. doi:10.1038/s41586-026-10836-6

76. Marvin AR, Marvin DJ, Lipkin PH, Law JK. Analysis of Social Communication Questionnaire (SCQ) Screening for Children Less Than Age 4. Curr Dev Disord Rep. 2017;4: 137–144. doi:10.1007/s40474-017-0122-1

77. Wolff JJ, Boyd BA, Elison JT. A quantitative measure of restricted and repetitive behaviors for early childhood. J Neurodev Disord. 2016;8: 27. doi:10.1186/s11689-016-9161-x

78. Exploratory Factor Analysis in R. Available: https://bookdown.org/luguben/EFA_in_R/

79. Revelle W. psych: Procedures for Psychological, Psychometric, and Personality Research. 2025. Available: https://cran.r-project.org/web/packages/psych/

80. Costa V, Sarmento R. Confirmatory Factor Analysis.

81. Genetic correlates of phenotypic heterogeneity in autism | Nature Genetics. [cited 17 Nov 2025]. Available: https://www.nature.com/articles/s41588-022-01072-5#Sec10

82. About SPARK for Autism - Autism Research. In: SPARK for Autism [Internet]. [cited 12 Nov 2025]. Available: https://sparkforautism.org/portal/page/about-spark/

83. TOPMed Imputation Server. [cited 21 Nov 2025]. Available: https://imputation.biodatacatalyst.nhlbi.nih.gov/#!

84. Mbatchou J, Barnard L, Backman J, Marcketta A, Kosmicki JA, Ziyatdinov A, et al. Computationally efficient whole-genome regression for quantitative and binary traits. Nat Genet. 2021;53: 1097–1103. doi:10.1038/s41588-021-00870-7

85. Robinson-Agramonte M de los A, Noris García E, Fraga Guerra J, Vega Hurtado Y, Antonucci N, Semprún-Hernández N, et al. Immune Dysregulation in Autism Spectrum Disorder: What Do We Know about It? Int J Mol Sci. 2022;23: 3033. doi:10.3390/ijms23063033

86. Zhou D, Gamazon ER. PsychENCODE models. Zenodo; 2020. doi:10.5281/zenodo.3859065

87. Zhou D, Jiang Y, Zhong X, Cox NJ, Liu C, Gamazon ER. A unified framework for joint-tissue transcriptome-wide association and Mendelian Randomization analysis. Nat Genet. 2020;52: 1239–1246. doi:10.1038/s41588-020-0706-2

88. Barbeira AN, Bonazzola R, Gamazon ER, Liang Y, Park Y, Kim-Hellmuth S, et al. Exploiting the GTEx resources to decipher the mechanisms at GWAS loci. Genome Biol. 2021;22: 49. doi:10.1186/s13059-020-02252-4

89. Wingo AP, Liu Y, Vattathil SM, Gerasimov ES, Mei Z, Ravindran SP, et al. Multiancestry brain pQTL fine-mapping and integration with genome-wide association studies of 21 neurologic and psychiatric conditions. Nat Genet. 2025;57: 2156–2165. doi:10.1038/s41588-025-02291-2

90. Gusev A, Ko A, Shi H, Bhatia G, Chung W, Penninx BWJH, et al. Integrative approaches for large-scale transcriptome-wide association studies. Nat Genet. 2016;48: 245–252. doi:10.1038/ng.3506

91. Foguet C, Gil L, Xu Y, Salazar-Magaña S, Ritchie SC, Persyn E, et al. OmicsPred as a centralised resource for genetic prediction of multi-omic traits. medRxiv; 2026. p. 2026.05.15.26353298. doi:10.64898/2026.05.15.26353298

92. Betti MJ, Lin P, Aldrich MC, Gamazon ER. Genetically regulated eRNA expression predicts chromatin contact frequency and reveals genetic mechanisms at GWAS loci. Nat Commun. 2025;16: 3193. doi:10.1038/s41467-025-58023-x

93. Zhang Z, Hong W, Ruan H, Jing Y, Li S, Liu Y, et al. HeRA: an atlas of enhancer RNAs across human tissues. Nucleic Acids Res. 2021;49: D932–D938. doi:10.1093/nar/gkaa940

94. Betti MJ, Gamazon E. eRNA GReX. Zenodo; 2024. doi:10.5281/zenodo.14027849

95. Yu G, Wang L-G, Han Y, He Q-Y. clusterProfiler: an R package for comparing biological themes among gene clusters. Omics J Integr Biol. 2012;16: 284–287. doi:10.1089/omi.2011.0118

96. Skene NG, Grant SGN. Identification of Vulnerable Cell Types in Major Brain Disorders Using Single Cell Transcriptomes and Expression Weighted Cell Type Enrichment. Front Neurosci. 2016;10. doi:10.3389/fnins.2016.00016

97. Siletti K, Hodge R, Mossi Albiach A, Lee KW, Ding S-L, Hu L, et al. Transcriptomic diversity of cell types across the adult human brain. Science. 2023;382: eadd7046. doi:10.1126/science.add7046

98. EWCE. In: Bioconductor [Internet]. [cited 1 Dec 2025]. Available: http://bioconductor.org/packages/EWCE/

